# Identification of Clonal Hematopoiesis Driver Mutations through In Silico Saturation Mutagenesis

**DOI:** 10.1101/2023.12.13.23299893

**Authors:** Santiago Demajo, Joan Enric Ramis-Zaldivar, Ferran Muiños, Miguel L Grau, Maria Andrianova, Núria López-Bigas, Abel González-Pérez

**Author notes:** Equal contribution (Santiago Demajo, Joan Enric Ramis-Zaldivar, Ferran Muiños).

## Abstract

Clonal hematopoiesis (CH) is a phenomenon of clonal expansion of hematopoietic stem cells driven by somatic mutations affecting certain genes. Recently, CH has been linked to the development of a number of hematologic malignancies, cardiovascular diseases and other conditions. Although the most frequently mutated CH driver genes have been identified, a systematic landscape of the mutations capable of initiating this phenomenon is still lacking. Here, we train high-quality machine-learning models for 12 of the most recurrent CH driver genes to identify their driver mutations. These models outperform an experimental base-editing approach and expert-curated rules based on prior knowledge of the function of these genes. Moreover, their application to identify CH driver mutations across almost half a million donors of the UK Biobank reproduces known associations between CH driver mutations and age, and the prevalence of several diseases and conditions. We thus propose that these models support the accurate identification of CH across healthy individuals

**Significance:** We developed and validated 12 gene-specific machine learning models to identify CH driver mutations, showing their advantage with respect to expert-curated rules. These models can support the identification and clinical interpretation of CH mutations in newly sequenced individuals.

## Introduction

In healthy hematopoiesis, a pool of hematopoietic stem cells (HSCs) contributes to all blood-related lineages. During aging, this process frequently gives place to clonal hematopoiesis (CH), a state in which one stem cell-derived population occupies a large fraction of the blood cells and platelets (1–6). This phenomenon of clonal expansion is driven by somatic mutations acquired by the HSCs at some point during life and is highly prevalent in elderly population (1–3,5–13). These mutations that affect an array of genes collectively named CH drivers, confer the HSCs bearing them growth advantages with respect to their neighbors, and are thus, under positive selection in hematopoiesis.

When the expanded clone presents with a variant allele frequency greater than 2%, and in the absence of any hematological disease phenotype, it is clinically referred to as CHIP, or clonal hematopoiesis of indeterminate potential (14). Although this entity most often presents without any clinical manifestation, epidemiological and molecular studies have, in the past decade, linked the presence of CH (above or below this clinical threshold) with an increased risk of development of myeloid malignancies, cardiovascular-related diseases, and all-cause mortality (2,6,7,14–18). Recent studies have also linked the presence of CH with the ulterior development of different conditions, such as solid malignancies (19), or with the response to certain infectious diseases (20).

In the three decades since the discovery of the genetic basis of CH, intensive research has led to the identification of some 60 of its driver genes (1,12,13). Nevertheless, we have, at best, a fragmented picture of which mutations of these genes have indeed the potential to drive this process of clonal expansion. While the ability of a few of them to drive CH has been tested in experimental models (21), replicating this for all possible mutations of CH genes constitutes a daunting experimental endeavor. The lack of a complete repertoire of CH driver mutations further complicates epidemiological studies of this phenotype and its relationship with other conditions. Across large cohorts of blood donors from the general population (sequenced at relatively low depth), such as the UK Biobank (22) or TOPMed (23), the ability to detect true CH driver mutations is hindered by the contamination with germline variants and sequencing artifacts (24). Thus, uncovering the repertoire of CH driver mutations is key, not only to understanding the molecular mechanisms underpinning CH, but also to accurately identifying individuals with CH and thus powering epidemiological studies of its relationship with more serious health conditions. This will also allow us to monitor the potential impact of CH driver mutations on the health of individuals carrying them.

Faced with this reality, several research groups and health-related international institutions have summarized the knowledge accumulated on several CH genes in a series of expert-curated rules to select the mutations most likely to drive CH (25–27). The application of these expert-curated rules, following a succession of stringent filters to the variants identified in the blood samples of healthy individuals, is a frequently used approach, among researchers, to identify CH cases (24).

Despite their practical usefulness, expert-curated rules have a series of caveats. First, their coverage of genes is heterogeneous, due to the difference in the amount of knowledge available for them or even across different domains of one gene. Secondly, expert-curated rules cannot be learned –or systematically updated– directly from information on CH mutations; rather this information must be first sedimented into shared knowledge. Thirdly, they can only be established for a small set of genes that have been intensely studied. We reasoned that these hurdles could be overcome by applying a machine learning-based approach that produces explainable models, trained on high-quality available CH mutations. This method would not suffer from any biases from sedimented knowledge, its models could reveal complex patterns in CH mutations that have not been apparent so far to expert knowledge and would be easy to scale as more datasets of CH mutations become available. This continuous expansion of explainable machine learning models of CH will increase our knowledge of the molecular mechanisms underlying it.

We reasoned that the approach we recently presented to build machine learning models inspired by evolutionary biology to distinguish cancer driver and passenger mutations in cancer genes (28) –using mutations observed across cancer patients– could be repurposed to produce CH-specific models. Using *bona fide* CH mutations identified across known CH driver genes (12) and sets of neutral hematopoiesis mutations, we trained machine learning models to identify CH driver mutations across 12 genes (collectively referred to as boostDM-CH). When tested on CH mutations not included in the training, the models show a performance that is in general terms above that of expert-curated rules. Models also outperform the results of experimental base-editing approaches. When applied to mutations identified in the blood of 470,000 donors from the general population (UK Biobank), the mutations identified by boostDM-CH models as CH drivers show a very significant association with age and the development of hematopoietic malignancies and other diseases, while mutations identified as CH non-drivers show no meaningful associations.

## Results

### BoostDM-CH models accurately identify CH driver mutations

Training machine learning models aimed at distinguishing CH driver mutations requires a high-quality dataset of blood somatic mutations identified across individuals. In principle, these could be obtained from blood sequencing datasets across large cohorts carried out in recent years (22,29). However, the set of somatic mutations identified by these single-sample low depth (usually ~30X) sequencing analyses is expected to be contaminated with germline variants and sequencing and mapping errors. Large cohorts of solid malignancies from different organs have been sequenced in the past 15 years, where one germline reference sample (usually blood) was used to call somatic tumor mutations. We reasoned that such cohorts could be repurposed to produce a set of high-quality blood somatic mutations using the tumor sample as germline reference (12). Following this idea, we identified blood single somatic nucleotide variants (SNVs) and indels (collectively, mutations) across more than 36,000 cancer patients from 3 large studies (TCGA, HMF, MSK-IMPACT) (30–32) (Fig. 1a). We call this process reverse mutation calling, as it reverses the normal use of these two samples in cancer genomics by using the tumor sample as the germline reference.

**Figure 1.**
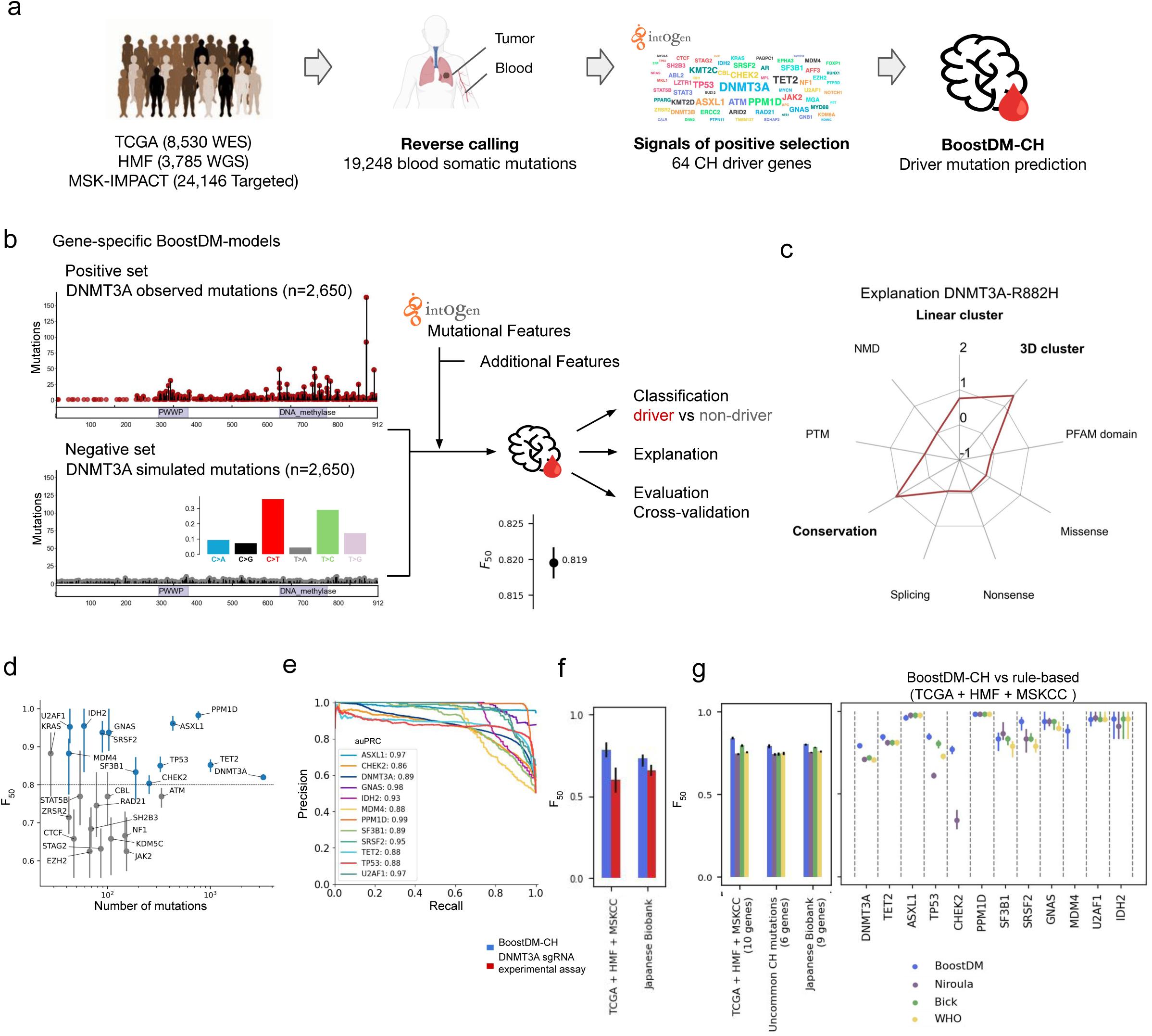
Building and evaluating BoostDM-CH models. a) Blood somatic mutations used to train the boostDM-CH models were identified in the three discovery cohorts through reverse calling (blood vs tumor sample); then, CH driver genes were identified by uncovering signals of positive selection using the IntOGen pipeline; finally, models to identify CH driver mutations were built. b) Training and cross-validation of machine learning-based boostDM-CH models, exemplified by *DNMT3A*. In the outcome of the model, mutations with a score equal to or above 0.5 are deemed CH drivers. c) Explanation of the classification of the *DNMT3A*-R882H mutation as a driver based on the contribution of the features employed in training the model. The numbers in the radar plot correspond to the SHAP values (34) of each feature. Features with positive SHAP value (i.e., positive contribution to the classification of a mutation as driver) appear above the ‘0’ line in the radar plot. d) Performance (median ± interquartile range (IQR) F_50_) of the cross-validation of 25 CH models as a function of their number of observed mutations. Blue dots represent the genes with high-quality models, i.e. F_50_ above 0.8 and sufficient discovery index to deem the set of mutations across the three discovery cohorts representative of their CH driver mutations. e) Area under the Precision-Recall curves of the 12 high-quality boostDM-CH models. f) Performance (median ± interquartile range (IQR) F_50_) of the classification of *DNMT3A* blood somatic mutations of boostDM-CH models and a *DNMT3A* experimental base editing assay. g) Performance (median ± interquartile range (IQR) F_50_) of the classification of blood somatic mutations of boostDM-CH models and three sets of expert-curated rules. Left, overall performance in three CH datasets; right, gene-specific performance in one of the datasets. PTM: post-translational modifications; NMD: nonsense-mediated decay.

Using the blood somatic mutations in these three discovery cohorts we previously identified 64 genes involved in CH development (12). We did this through the detection of signals of positive selection in their pattern of mutations across samples using a toolbox of methods originally developed to identify cancer driver genes, organized in the Integrative OncoGenomics (IntOGen) pipeline (intogen.org) (33) (Fig. 1a). We then aimed to build gene-specific machine learning models that captured combinations of features (mostly computed through the methods in the IntOGen pipeline) characteristic of the CH driver mutations found in each gene. These features include, for example, the significant clustering of mutations in specific regions of the linear sequence or the three-dimensional folded structure of the protein, the enrichment of mutations in certain domains, the consequence type of each mutation, and some additional features such as the conservation of the residue and post-translational modifications (PTM) (28).

The most important and challenging step to train these models is to start with a good quality set of driver/positive, non-driver/negative blood mutations. Using a set of known CH driver mutations as the positive set could bias the models toward the current knowledge. To solve this problem we reasoned –as we did previously in the case of cancer– that somatic mutations detected in human blood, an unbiased set enriched for driver mutations of each CH gene, constitute the best positive training set. The ideal negative set contains mutations that could have occurred by neutral mutagenesis processes in HSCs but do not provide a selective advantage (non-drivers). We thus generated synthetic mutations by simulating neutral mutagenesis in HSCs, following the probabilities of tri-nucleotide changes observed across blood samples as the negative training set. Although these sets are highly enriched in driver and non-driver mutations respectively, they are imperfect, as the group of observed CH mutations may contain non-drivers, whereas the synthetic set of mutations may contain drivers. These imperfections need to be taken into account in the strategy used to train the models.

For example, to build a model for DNMT3A, we used the 2,650 blood somatic mutations observed across the donors of the three discovery cohorts as the positive set (Fig. 1b, left top panel), and created fifty negative sets of 2,650 synthetic mutations (Fig. 1b, left bottom panel). Then, we trained fifty base (extreme gradient boosting) classifiers on randomly chosen 70% of the positive set and the same fraction of each of the negative sets. This is designed specifically to deal with the expected imperfection of the positive and negative sets of mutations. Finally, these fifty base classifiers integrated a full DNMT3A model, and mutations held back from the training (30% in each base classifier) and evaluated as a test set of each of the base classifiers provided a cross-validation for this full model. Since some of the mutations in the positive set may be CH non-drivers, we selected the Fscore-50 (F_50_) –a metric that gives more weight to precision than recall– to evaluate the performance of the integrated model. The DNMT3A model exhibits an F_50_ of 0.82 upon cross-validation.

We aimed to produce explainable models capable of capturing complex relationships between biological features that may be unattainable to expert-curated rules (see Introduction). Therefore, we decomposed the prediction cast for every mutation to reveal the contribution of individual features, using the Shapley additive values (SHAP) strategy (34). For example, for the recurrent R882H mutation of DNMT3A (Fig. 1c), the prediction as CH driver results primarily from a combination of three salient features: its location within a linear (35) or a three-dimensional (36) cluster and its conservation across vertebrate species (37).

We followed the same strategy for the 25 CH driver genes with sufficient number of mutations to carry out the training (Methods). We refer to these full models, collectively, as boostDM-CH (extreme gradient boosting of CH driver mutations). We also computed a discovery index for each of these genes to assess how well the mutations observed across the three discovery cohorts represent all potential CH driver mutations in these genes. This discovery index tracks the increase in the number of observed mutations in a gene as more blood samples are sequenced (28). The boostDM-CH models of 12 CH driver genes (*ASXL1*, *CHEK2*, *DNMT3A*, *GNAS*, *IDH2*, *PPM1D*, *SF3B1*, *SRSF2*, *TET2*, *TP53*, *U2AF1*, and *MDM4*) yielded an F_50_ above 0.80 and their discovery indexes were deemed sufficiently representative of all their potential driver mutations (genes in blue in Fig. 1d; Fig. S1; Methods and Supplement). The *KRAS* model, although scoring an F_50_ above 0.8 does not satisfy the threshold of discovery index (Methods). As a trend, genes with more observed CH mutations (or higher discovery index) yielded models with higher F_50_ (Fig. 1d and S2a). Nevertheless, some CH driver genes with relatively low number of mutations observed across the three discovery cohorts, most of them concentrated within significant clusters (e.g., *U2AF1* and *IDH2*), also yielded high-quality models. The area under the cross-validation precision-recall curves (AUC) for these models ranged between 0.87 and 0.99 (Fig. 1e; Fig. S2b).

To benchmark the performance of the boostDM-CH model of *DNMT3A*, the most recurrent CH driver gene, we compared it with a recently published experimental base editing assay that quantified the reporter methylation activity of mutants (38). We reasoned that the best way to measure the performance of any experiment or *in silico* method in the identification of CH driver mutations is to compare them to the somatic mutations observed in CH and synthetic mutations generated following the trinucleotide frequencies observed in normal hematopoiesis, as we did previously in the case of cancer drivers (28). As explained above, these two sets are enriched for CH drivers and non-drivers, respectively. First, we compared the classification of CH mutations carried out in the cross-validation of the *DNMT3A* model (consisting, as explained above, of mutations left out of the training of each base model) with the base editing assay of the gene that assessed the loss in methylation capability of the mutants. The *DNMT3A* model showed a better performance in its cross-validation than this experimental assay in the separation between observed and neutral CH mutations (Fig. 1f, left bars; Fig. S2c). Moreover, when applied to an independent set of CH mutations (i.e., not the ones from the discovery cohorts) identified across the general population (Japanese Biobank) (29), it also showed higher F_50_ than the experimental assay (Fig. 1f, right bars; Fig. S2c).

We then compared the collective performance of boostDM-CH models to that of three sets of expert-curated rules (referred to as Niroula (25), Bick (26), and WHO (27), Table S1) on three different sets of CH mutations. We first compared the performance of boostDM-CH models and the expert-curated rules on the classification of cross-validation CH observed and synthetic neutral mutations from the three discovery cohorts across 10 CH genes (*MDM4* and *CHEK2* are not covered by the three rule sets; Fig. 1g left bars; Fig. S2d). Next, we compared the performance of boostDM-CH models with expert-curated rules in the classification of a set of rare CH mutations in 6 genes (10), included neither in the training nor in the cross-validation of models (center bars). Finally, we compared boostDM-CH models and the expert-curated rules in the identification of CH mutations across the Japanese Biobank (right bars). Interestingly, the performance of boostDM-CH models is systematically greater than that of the three sets of expert-curated rules, even when analyzing mutations not employed in their training. This performance comparison reflects a variability across the 10 CH drivers genes (Fig. 1g; Fig. S2d). These results demonstrate the potential of machine-learning models trained from observed mutations –and thus, unbiased by prior knowledge– to capture the genetic mechanisms underlying CH.

### The repertoire of CH driver mutations reveals mechanisms underlying CH

We next used boostDM-CH models to classify all possible mutations in each CH driver gene (*in silico* saturation mutagenesis). Each mutation is classified on the basis of the boostDM-CH score resulting from the aggregation of trees into each base classifier: mutations attaining a score equal to or above 0.5 are classified as CH drivers, while those with score lower than 0.5 are deemed CH non-drivers (Fig. 2a,b). While many driver mutations exhibit boostDM-CH scores very close to 1 (agreement between the majority or all trees within base classifiers), some score lower, even close to the 0.5 boundary.

**Figure 2.**
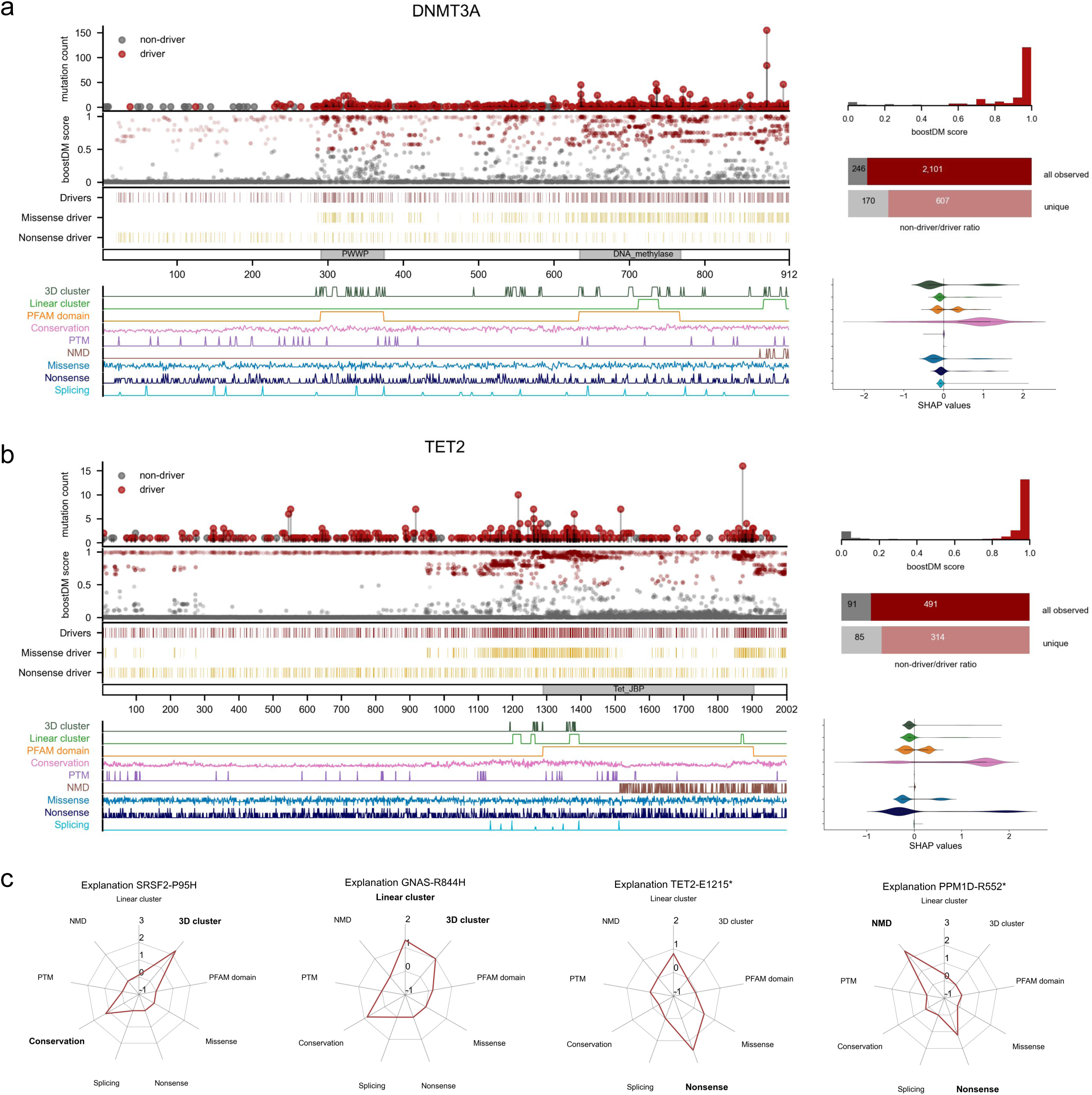
In Silico Saturation Mutagenesis of CH genes. Blueprints of CH driver mutations in *DNMT3A* (a) and *TET2* (b). The plots represent the distribution of driver and non-driver mutations, with red representing predicted driver mutations (boostDM-CH score >= 0.5) and gray, non-driver mutations. From top to bottom, the first plot contains observed mutations in the three training cohorts classified as drivers or non-drivers, with the height of each needle tracking the recurrence of the mutation. The distribution of the boostDM scores of these mutations observed in blood samples is shown at the right, with the partition of all (top) or unique (bottom) observed mutations shown in bars. The plot immediately below the observed mutations presents boostDM-CH scores of all possible SNVs along the protein sequence. The density of all potential SNVs classified as drivers is presented immediately below, with a distinction between missense and nonsense drivers (gold). The values of mutational features used to train the models are shown linearly along the protein sequence in the plot at the bottom of the figure. The violin plots at the right present the distribution of SHAP values of all driver mutations across the features used to train the models. The SHAP values entail a decomposition of the boostDM score of mutations into the contribution of each of these features. c) Radar plots representing the SHAP values for several illustrative driver mutations.

The distribution of driver CH missense and nonsense mutations in *DNMT3A* differ, with the former enriched at specific stretches of the gene, and the latter more smoothly distributed along its sequence (Fig. 2a). The clustering of observed mutations at specific regions of the protein 3D structure (and/or linear sequence stretches) and at the PWWP and DNA methylase domains appear the most important contributors to the identification of driver missense mutations (Fig. 2a). Possibly, these clusters of missense driver mutations represent interference with different aspects of the *DNMT3A* methylation activity. For example, while R882 mutations are known to affect its tetramerization, thus acting as a dominant negative (5), amino acid substitutions in the PWWP domain interfere with the recognition of histone modifications (38,39). Conversely, nonsense mutations that trigger nonsense-mediated decay would result in a reduction of *DNMT3A* abundance in the cell.

*TET2* missense CH driver mutations (some of which are known to act in haploinsufficiency) (40) appear enriched for certain regions of the protein, grouped in three-dimensional and linear clusters, whereas nonsense mutations appear uniformly distributed along its entire sequence (Fig. 2b). The clusters of CH driver missense mutations overlapping different stretches of the protein, most of them within the Tet domain or in its proximal region, may be related to distinct aspects of the catalysis of the methyl group removal from CpGs (41); these mutations may result in anomalies of the methylation profile during hematopoiesis (42).

The analysis of the distribution of CH driver mutations across observed blood somatic mutations or all possible mutations reveals different landscapes within the remaining 10 CH driver genes with boostDM-CH models (Figs. S3 and S4). A group of CH drivers (such as *ASXL1*, *TP53*, or *CHEK2*) appears to act in a loss-of-function manner, showing a broad distribution of missense and nonsense CH driver mutations along long tracts of the protein sequence. This contrasts with another group of genes (*SF3B1*, *PPM1D*, *U2AF1*, and *IDH2*) known to act in a gain-of-function manner, where CH driver mutations appear confined to specific regions of the protein, and in extreme cases, to a single mutational hotspot. These clusters of CH driver mutations are related to underlying alterations of the biological function of these genes.

A detailed exploration of the contribution of different features to the classification of four CH driver mutations across the same number of genes is presented through the radial plots in Figure 2c. For example, in the case of *PPM1D*, nonsense CH driver mutations tend to concentrate towards the C-terminal portion of the protein. These mutations escape nonsense-mediated decay and result in a form of the protein lacking a degron sequence and, thus, with increased stability (43) (Fig. 2c). This, in turn, causes altered phosphorylation of proteins involved in the response to DNA damage, such as *TP53*, providing mutant cells with an advantage when exposed to certain cytotoxic therapies (11,43).

The boostDM-CH models of these genes encapsulate the knowledge of the molecular mechanisms of CH by directly exploiting the signals of positive selection presented by their observed mutational patterns when compared to that expected under neutrality. Certain differences are apparent between the configuration of CH driver mutations in these genes according to boostDM-CH models and expert-curated rules (Fig. S5). There are also similarities and differences in the configuration of CH and cancer driver mutations in 3 of these CH driver genes for which we have also been able to build boostDM cancer models (28) (Fig. 3a-c and Fig. S6). One of the differences corresponds to a mutational hotspot of *IDH2* observed in myeloid malignancies, but absent in CH mutations (Fig. 3c). This hotspot is considered as a driver CH mutation by the expert-curated rules, which include data from hematological malignancies. However, its absence from the output of the *IDH2* model suggests it may be an example of incorrect definition of the rule. This is an example of the benefits provided by models trained from observed data over expert-curated rules established from sedimented knowledge.

**Figure 3.**
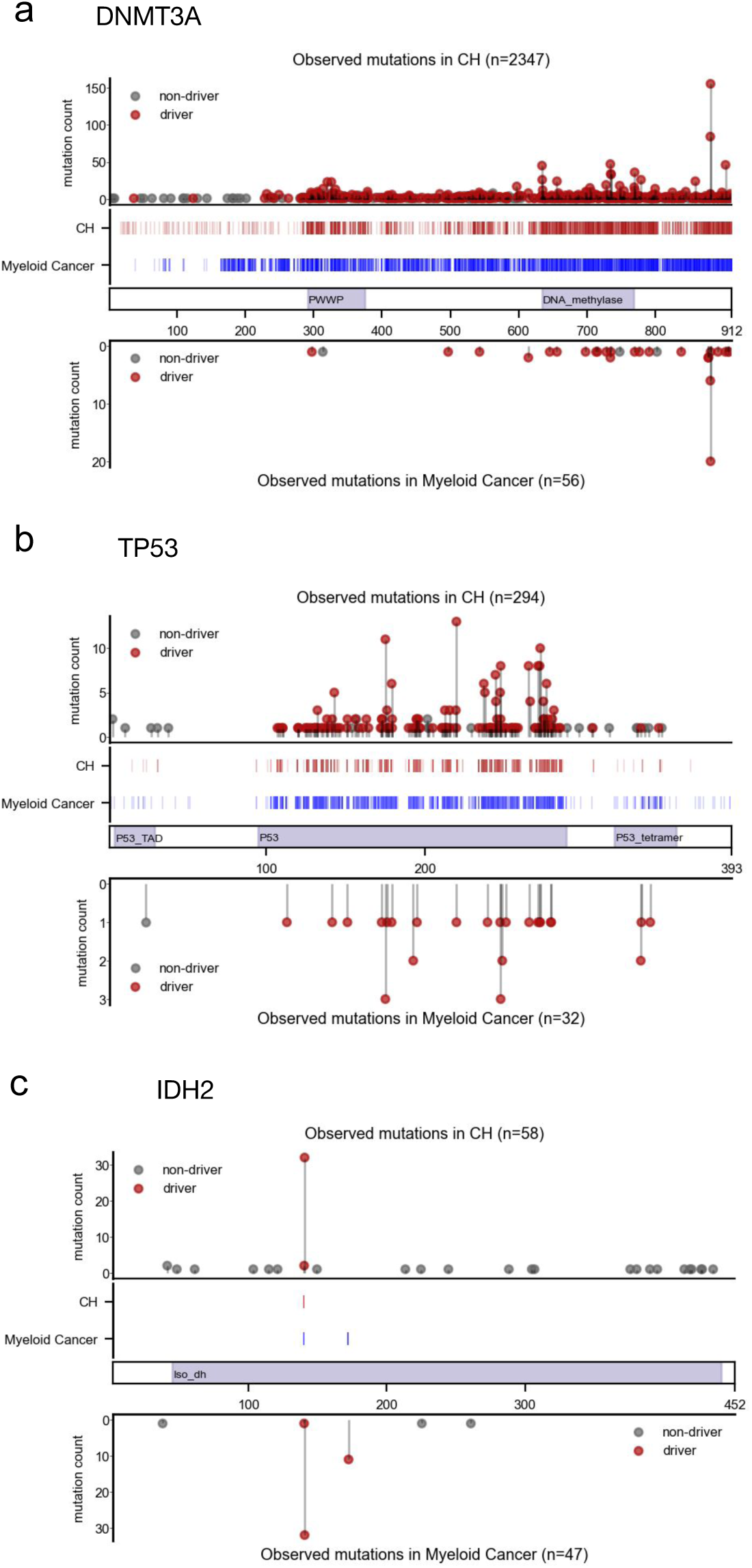
Comparison of boostDM-CH models and myeloid boostDM models in three genes. For each gene (a-c), the needle plots represent the distribution of driver and non-driver observed mutations in CH (top) and myeloid cancer (bottom) cohorts. The internal plots represent the distribution of driver mutation along the sequence of each gene using *in silico* saturation mutagenesis by boostDM, in CH (red) and myeloid cancer (blue).

In summary, BoostDM-CH models support the exploration of underlying CH mechanisms across genes. To facilitate this process, the results of the *in silico* saturation mutagenesis provided by boostDM-CH models of the 12 CH driver genes included in this section are available to the research community at www.intogen.org/ch/boostdm.

### BoostDM-CH models identify CH driver mutations in a large general population cohort

One of the main hurdles to exploiting large cohorts of donors (such as the UK Biobank, UKB) for population-wide CH epidemiological studies is the difficulty to accurately identify blood somatic mutations (in a relatively shallow sequencing) without a reference germline sample from the same donor (12,24). Potential somatic mutations identified across these blood samples may contain a non-negligible fraction of germline variants and sequencing artifacts, as well as passenger mutations. We reasoned that boostDM-CH models –trained on high-quality blood somatic mutations obtained from the reverse mutation calling across tumor cohorts– could be employed to accurately identify CH driver mutations in this setting. We could then use these CH driver mutations to analyze the relationship between CH and several phenotypes across the population.

Thus, we next identified 201,916 potential somatic variants in the 12 genes with high-quality boostDM-CH models across the blood samples of 467,202 individuals in the UKB (Methods). BoostDM-CH models were employed to sift these potential somatic variants. We identified 41,311 CH driver mutations (28,508, or 69.0%, non-synonymous; 10,098, or 29.3%, nonsense; and 705, 1.7%, splice site affecting) in the 12 genes (Fig. 4a; Methods). CH driver mutations in these 12 genes appeared in 8.2% (38,129) of the donors of the UKB cohort (92.5% of them bearing a single driver mutation). While all potential somatic variants exhibited a marked bimodal distribution of variant allele frequency (VAF) with one mode very close to zero, and a second mode around 0.5, CH driver mutations show a salient peak at very low VAF, with a small tail that extends up to 0.5 (Fig. 4b; Fig. S7a). This reflects that the approximately 165,000 SNVs identified by boostDM-CH models as non-drivers probably comprise a mixture of germline variants, sequencing artifacts, and passenger somatic mutations. This is also implied by the differences in the tri-nucleotide mutational profiles of variants identified as CH drivers or non-drivers by boostDM-CH (Fig. S7b). Most of the variants identified as drivers possess boostDM-CH scores above 0.9, while the overwhelming majority of non-drivers are scored below 0.1, forming an extremely bimodal distribution (Fig. S7c,d). Taking advantage of this, we can define more restrictive sets of high-confidence drivers and non-drivers using these two thresholds (Fig. S7).

**Figure 4.**
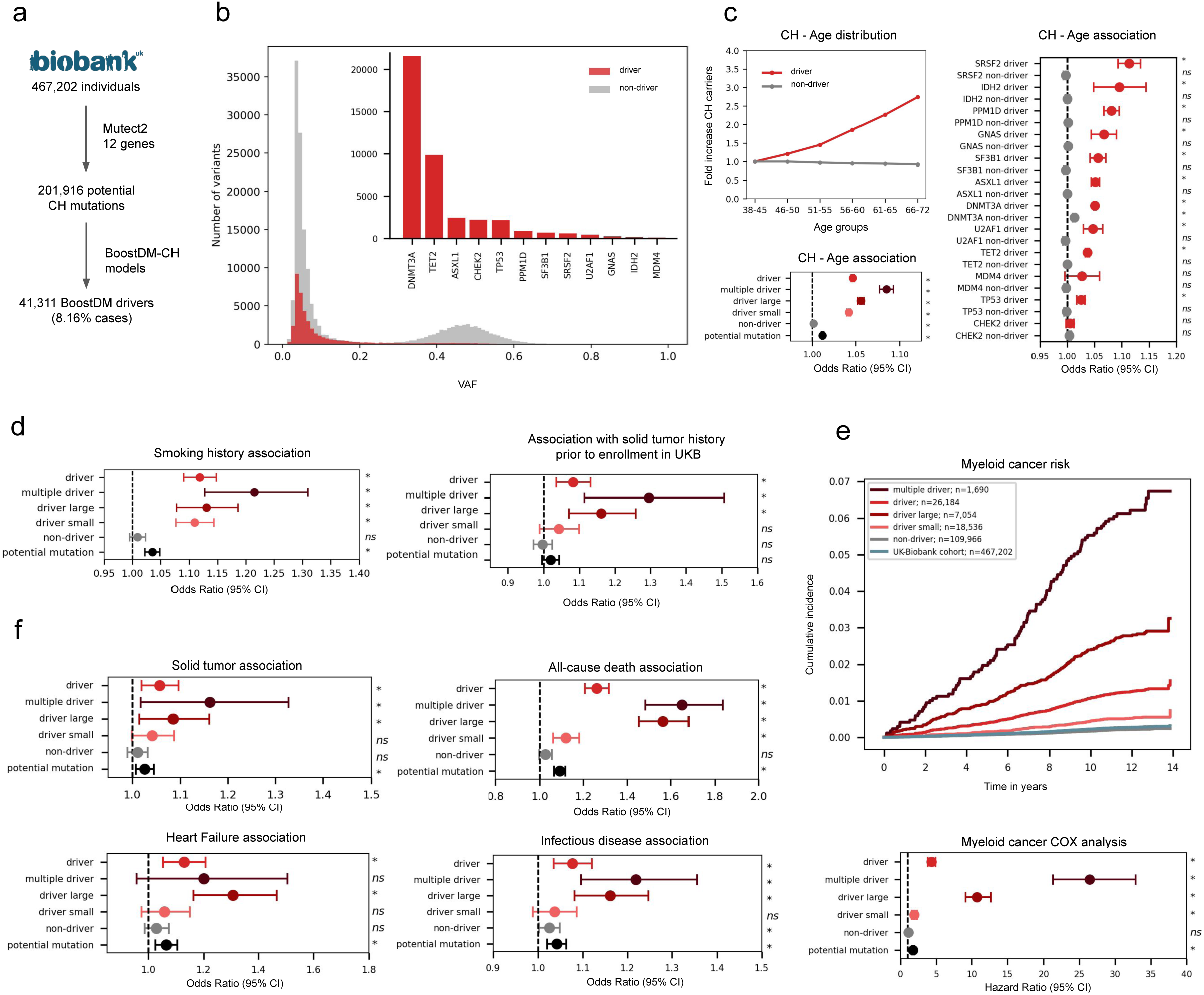
Application of BoostDM-CH to identify CH driver mutations across 467,202 donors. a) Identification of CH driver mutations in UKB donors using boostDM-CH models. b) Distribution of VAF of CH driver and non-driver mutations; inner, number of driver mutations identified in the 12 CH driver genes studied. c) Fold increase in the proportion of cases with CH driver and non-driver mutations across age groups (upper left). Significance of the overall (bottom left) or gene-wise (right) association with age measured via logistic regression. In this and subsequent regression plots, several sets of donors in the UKB are selected for analysis: driver, donors bearing at least one CH driver mutation (according to boostDM-CH); multiple driver, donors bearing more than one CH driver mutation; driver large, donors bearing at least one CH driver mutation with VAF >= 10%; driver small, donors bearing aCH driver mutation(s) with VAF < 10%; non-driver, donors bearing a potential mutation in a CH driver gene classified as non-driver by boostDM-CH; potential mutation, donors bearing any potential mutation (driver or non-driver) in a CH driver gene (see Methods). d) Association of CH with known causative factors including smoking and solid tumor history (as proxy of chemotherapy) prior to enrollment in UKB as a proxy of treatment, measured via logistic regression. e) Association of CH with the risk of myeloid malignancies, measured via Kaplan-Maier analysis (upper) and Cox proportional hazards model (bottom). f) Association of CH with all-cause death, heart failure, the occurrence of a solid tumor, and any infection (measured via a composite variable). All logistic regressions and Cox analyses included age, sex, and 10 ancestry principal components as covariables. Additionally, tumor-related associations also included smoking history as covariates. Heart failure associations included smoking history, dyslipidemia, body mass index, hypertension, and diabetes type II status as covariates whereas Infectious diseases included smoking history and occurrence of hematological neoplasm. Asterisks represent significant associations (FDR<0.05); ns: non-significant.

The proportion of individuals with CH driver mutations identified by boostDM-CH models (relative to those in the youngest age bracket, 38-45 years old) grows with age, as expected of true CH mutations (Fig. 4c). In contrast, the number of donors with non-driver CH mutations remains constant. While all potential somatic SNVs in the 12 CH driver genes show a significant association with age (Fig. 4c, black dot), the significance is virtually entirely due to CH driver mutations (top red dot), since non-driver mutations (gray dot) show almost no association with age. The effect size is larger for individuals with CH mutations with VAF > 10% (driver large) than for those with CH mutations with VAF <= 10% (driver small), and it also appears larger for donors bearing several co-occurring CH driver mutations (multiple drivers). This difference in the association with age is even clearer when high-confidence CH driver and non-driver mutations are compared (Fig. S7e). CH driver mutations in individual genes show higher association with age than non-driver mutations, although for some genes this association is not significant due to the small number of mutations observed in the cohort (Fig. 4c). In contrast, non-driver mutations in nearly all genes show no significant association with age.

We next verified known associations of CH, defined by the presence of CH driver mutations identified by boostDM-CH, with the exposure to external CH promoters across UKB individuals. Thus, we corroborated that individuals with a history of smoking have significantly higher likelihood of carrying CH driver mutations than non-smokers, as have donors who suffered a solid malignancy (as a proxy of exposure to cytotoxic drugs) (1,12,13) prior to their enrollment in UKB (Fig. 4d, Fig. S8a,b), while we observed no association with CH non-drivers. We also checked the association of CH driver mutations identified by boostDM-CH in UKB with subsequent conditions known to be linked to CH. First, we corroborated the known association of the presence of CH driver mutations with the subsequent development of hematopoietic malignancies, in particular of myeloid origin (Fig. 4e; Fig. S9a-f). Both, the significance of the association and the increase in risk appear higher for donors bearing high-confidence CH drivers (Fig. S9f), large CH clones, or clones with multiple mutations (Fig. 4e). Conversely, the risk of donors bearing CH non-driver mutations to subsequently develop any type of hematopoietic malignancy is comparable to that of the general UKB population. We also verified that the known associations between CH driver mutations and increased risk of subsequent heart failure, development of a solid malignancy, risk of any type of infection, and all-cause mortality are reproduced (Fig. 4f, S10-S12). Importantly, all aforementioned associations appear as significant –or slightly more significant– with CH driver mutations identified by boostDM-CH than selected using expert-curated rules established by CH researchers (Fig. S13 and S14).

### The fitness of CH driver mutations

What distinguishes driver and non-driver CH mutations is that the former provide an increase of fitness to the HSCs where they occur, thus resulting in its expansion in blood. To estimate the fitness provided by specific mutations, we exploited the distribution of their VAF across UKB individuals (44). In detail, we computed the gain in fitness from the VAF distribution of 220 SNVs that were recurrently observed (in at least 30 cases) across UKB donors, after filtering out potential germline variants and variants with multimodal VAF distribution (Methods, Fig. S15). Of these, 136 were classified as CH drivers by boostDM-CH, while the remaining 84 were classified as CH non-drivers (Fig. 5a). As expected, the fitness estimated for CH driver mutations was significantly higher than that of non-drivers (Fig. 5b), a trend maintained across genes (Fig. 5c). In agreement with previous findings, we observed that driver CH mutations affecting splicing factors were among those conferring the highest fitness (44,45).

**Figure 5.**
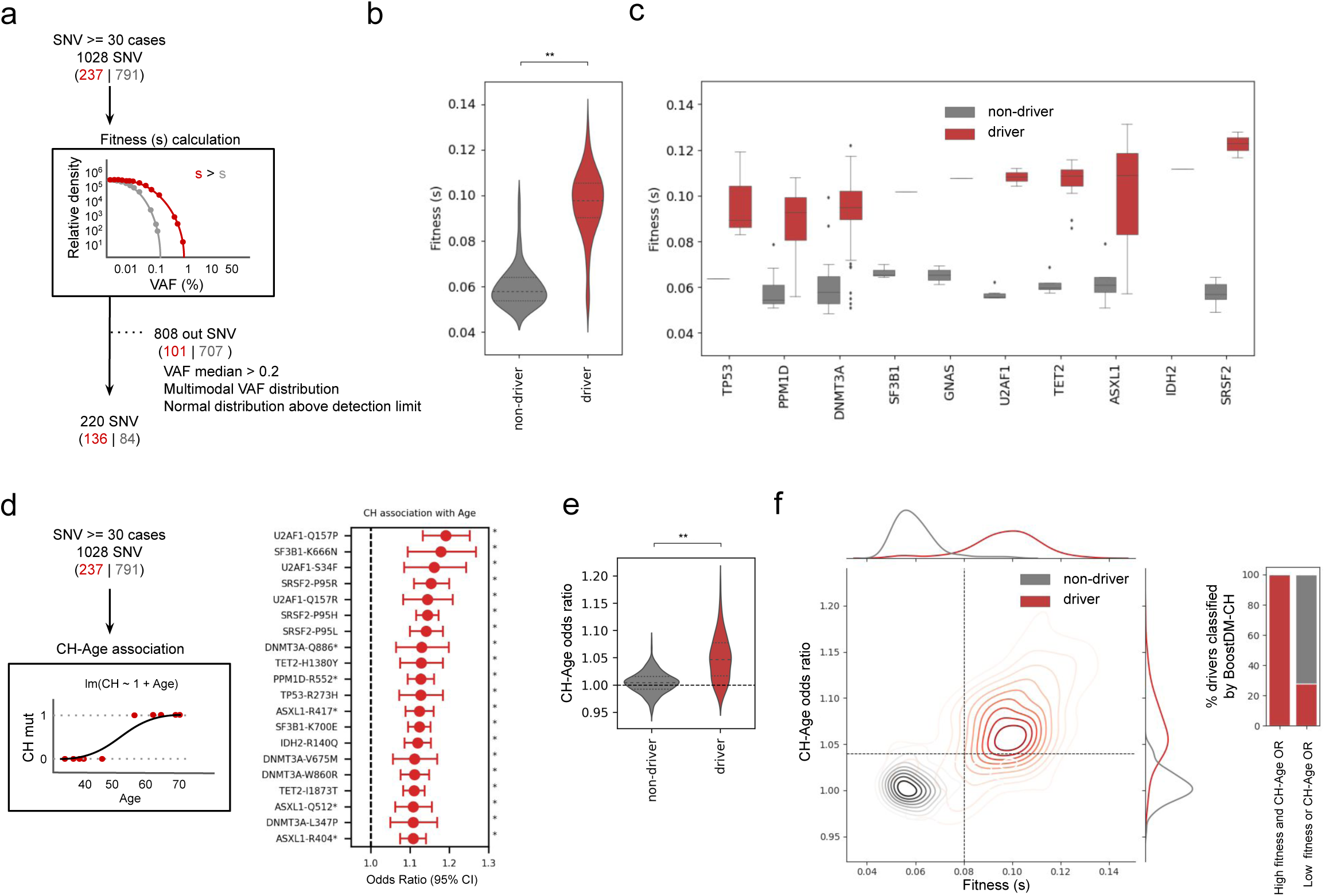
The fitness of recurrent blood somatic SNVs across 467,202 UKB donors. a) Schematic representation of the fitness score calculation for individual recurrent blood somatic SNVs, based on their observed VAF distribution across the UKB cohort. Red refers to drivers and grey for non-drivers. b,c) Overall (b) and per gene (c) distribution of fitness scores of CH driver and non-driver mutations. d) Left, schematic representation of the calculation of the associations (odds-ratio and p-value) between individual CH driver SNVs and age using logistic regression; right, top 20 SNVs with higher association with age. Red refers to drivers and grey for non-drivers. e) Distribution of odds-ratios computed for the association of driver and non-driver CH mutations with age (ageOR). f) Left, kernel density estimate (KDE) representing the bi-dimensional density of SNVs in the ageOR-fitness score plane. The horizontal dashed line represents an ageOR of 1.04, the minimum rendering a significant p-value, whereas the vertical line is arbitrarily set at a fitness score of 0.08. These lines effectively separate the bi-dimensional distributions of driver and non-driver recurrent CH SNVs into four quadrants, with the density of both distributions clearly separated in the top-right (drivers) and bottom-left (non-drivers) quadrants. The distribution of fitness and ageOR for driver and non-driver mutations are presented along the x-axis and the y-axis of the plot, respectively. The proportions of driver and non-driver mutations in these two quadrants of the plot are represented in the stacked bar plots at the right of the plot. In the figure, red denotes CH driver SNVs, whereas gray represents CH non-driver SNVs. Asterisks represent significant associations (FDR<0.05). Double asterisk represents p-value <0.001.

We also computed the strength of the association with age of each of these recurrent 220 variants (Fig. 5d) and found that, as expected, mutations classified as CH drivers by boostDM-CH appear significantly more associated with age than non-drivers, irrespective of their recurrence (Fig. 5e). In summary, among the most recurrent variants identified in 12 CH genes in UKB, those identified as drivers by boostDM-CH tend to confer high fitness to HSCs, and show an association with age. In contrast, recurrent non-driver variants in the same genes lack the combination of these two features (Fig. 5f). Furthermore, all recurrent mutations that confer high fitness and strong association with age (top right quadrant in Fig. 5f and first bar in the barplot at the right), are drivers according to boostDM-CH, as the chance to detect high fitness clones through sequencing increases with age. Conversely, only a minority of the recurrent mutations that confer low fitness and show no significant association with age (bottom left quadrant and second barplot) are identified as drivers. Restricting the analysis to samples with only one mutation in any of the 12 genes produced (Fig. S16) and to non-observed or non-recurrent mutations in the training set (Fig. S17a,b; Supp. Note) produced similar results.

In summary, a number of orthogonal pieces of evidence obtained from the population represented in the UKB, from the association with age and health risks to the estimated gain in fitness, demonstrate the effectiveness of using boostDM-CH to identify driver CH mutations.

## Discussion

Here we built machine learning-based models to identify all CH driver mutations in 12 well-known CH genes. We were primarily motivated by the lack of unbiased methods to identify the mutations responsible for the development of CH, even within well-established CH driver genes (24). One key requirement to train such machine learning models is the availability of a big enough set of high-quality blood somatic mutations. To fulfill this, we resorted to cohorts of cancer patients where blood somatic mutations can be reliably identified through a comparison of blood and tumor samples (12). The models were then trained on features that distinguish these blood somatic mutations observed in CH genes across donors from those expected to arise under neutral mutagenesis (28).

We demonstrate that these models perform on par with –or slightly better– than expert-curated rules designed by CH researchers on the basis of years of accumulated knowledge in the task of distinguishing newly observed CH mutations from neutrally arising blood mutations (25–27). Similar –or slightly better– performance with respect to these rules is observed when boostDM-CH models are applied to the task of distinguishing CH driver mutations in these genes from a combination of sequencing artifacts, germline contamination, and passenger somatic mutations across half a million donors in the UKB (22). This demonstrates that machine learning models trained unbiasedly on observed blood somatic mutations are able to recapitulate sedimented knowledge on CH development.

BoostDM-CH models can be employed to gain new insights into the mutational mechanisms underlying CH in different genes. Here, we provide examples with *DNMT3A*, *TET2*, and the models of other genes. The 12 boostDM-CH models trained in this work and their associated data are available to the CH research community at www.intogen.org/ch/boostdm. In the future, as more cohorts for which blood and a second sample (such as in cancer patients cohorts) become available in the public domain, growing sets of reliable blood somatic mutations will be identified, and good-quality models for more CH genes will be within reach.

Moreover, we have shown a path for the identification of CH driver mutations across donors in a large cohort, from whom only a blood sample is available, using boostDM-CH models. Applying this rationale to the UKB, we are able to recapitulate known associations of CH with age and the development of several phenotypes. We show that *ad hoc* laborious filtering steps of the variants detected in these blood samples can be easily replaced by the use of boostDM-CH models. We envision that this will streamline, accelerate and increase the reproducibility of studies that require reliable identification of CH driver mutations.

We have previously employed machine learning-based models trained using the same evolutionary-inspired approach described here to identify driver mutations in cancer genes (28). Here, we report for the first time the use of boostDM-like models (this time aimed at identifying CH driver mutations) to systematically identify CH across a large cohort of donors. We show that this leads to the re-discovery of well-established associations between life-style exposures and an increased risk of developing CH and between presence of CH and increase of the risk of subsequent conditions. This work constitutes a proof-of-principle for a wider use of boostDM-CH models in large retrospective or prospective clinical studies aimed at discovering such associations. Furthermore, it also illustrates how the models could assist in detecting CH across large populations, to subsequently monitor individuals at risk of developing different conditions.

In summary, in this article and our previous work on the discovery of CH driver genes (28), we provide a systematic approach to advance the study of CH through computational approaches. As larger cohorts of blood somatic mutations become available, boostDM-CH models for more genes will be within reach.

## Methods

### Blood somatic mutations in three discovery cohorts

Blood somatic mutations identified across two of our discovery cohorts –TCGA (whole-exome) (30), HMF (whole-genome) (31)– through reverse calling were obtained from our previous study Pich *et al*., 2022 (12). Briefly, aligned sequencing reads from normal (blood) and tumor BAM files from patients with solid tumors in TCGA and HMF cohorts were compared (using Strelka2) to call blood somatic mutations, which were subjected to a strict filtering post-process described in detail in Pich et al, 2022 (12). We thus retrieved all mutations identified in each of these two discovery cohorts. Blood somatic mutations in a third discovery cohort –MSK-IMPACT (in a panel of genes) (32) called by Bolton *et al*., 2020 (1)– were obtained from cBioPortal (https://www.cbioportal.org/) (46).

### Compendium of mutational CH genes

In the previous work by Pich *et al*., 2022 (12) mutations identified in the three discovery cohorts were used to detect genes under positive selection through the measurement of deviations from the mutational patterns expected under neutrality (12,33,35,36,47–51). This analysis resulted in 64 genes with signals of positive selection in CH in at least one of the three cohorts. These 64 genes were used as the starting point of the work described here. Collecting the somatic mutations identified in each of them in the three discovery cohorts, we attempted to build boostDM-CH models for all of them. We completed the process of training for the 25 genes with at least 30 mutations in the training set after the training-test partition of the base classifiers (see below). The CH driver genes finally included in this process were *ASXL1*, *ATM*, *CBL*, *CHEK2*, *CTCF*, *DNMT3A*, *EZH2*, *GNAS*, *IDH2*, *JAK2*, *KDM5C*, *KRAS*, *MDM4*, *NF1*, *PPM1D*, *RAD21*, *STAT5B*, *SF3B1*, *SH2B3*, *SRSF2*, *STAG2*, *TET2*, *TP53*, *U2AF1*, *ZRSR2*.

### BoostDM-CH models

BoostDM-CH delineates a supervised learning strategy based on observed mutations in sequenced blood samples and their site-by-site annotation with mutational features, comparing observed mutations in genes for which the consequence-type-specific excess is high enough (48) with randomly selected mutations following the trinucleotide change probability. The method essentially looks into the protein coding sequence of the genome as all mutations considered map to the canonical transcripts in protein-coding genes according to VEP v.101 (52). The training of boostDM-CH models (as the calculation of CH drivers discovery index and the *in silico* saturation mutagenesis) follows the rationale previously presented for cancer boostDM models (28). These steps for boostDM-CH models are described in detail in Supplementary Methods.

### SHAP local explanations

We use the Shapley additive values strategy (SHAP) (34,53) as a means to obtain an additive decomposition of the boostDM-CH score in the space of features used to train a model (28). Each of the 50 base classifiers integrated in the boostDM-CH model of a gene (see Supplementary Methods) yields an additive explanation model based on SHAP. Specifically, for each base classifier we can decompose the logit forecast yielded for a particular mutation into a vector of SHAP values, one per feature, in such a way that their sum is equal to the logit predicted probability for that mutation. In order to provide a consensus SHAP value for each feature and mutation across base classifiers, we simply take the average of the 50 values.

### Implementation

The boostDM-CH pipeline has been implemented in Python and Nextflow (54). The base classifiers were trained with XGboost (55) v.0.90 and we used SHAP (34,53) v.0.28.5 to compute the local explanations for the predictions of the base classifiers. The pipeline is available as a GitHub repository: https://github.com/bbglab/boostdm-pipeline/tree/ch.

### In silico saturation mutagenesis

In silico saturation mutagenesis is a term commonly used to indicate the assessment of all possible changes in a gene or protein with a computational approach (28). To assess the driver potential of all possible mutations in CH driver genes, we used boostDM-CH models.

### Benchmarking boostDM-CH models

In order to validate the *in silico* saturation mutagenesis approach implemented via boostDM-CH models we compared the cross-validation performance (F_50_) of boostDM-CH models with two other approaches: 1) a simple classifier (logistic regression) derived from the readouts of an experimental base editing assay of *DNMT3A* (38) and 2) three commonly used sets of expert-curated rules (25–27). For these analyses, we used three different datasets of mutations.

### Datasets used for validation

#### Observed mutations across discovery cohorts

We built two validation datasets using the cross-validation splits used to train the 50 base classifiers of boostDM-CH, which we can deem “permissive” and “stringent”, respectively. In both cases we collected all the mutations found across the test splits of each of the 50 base classifiers (see Training Splits section above) with their true labels and the prediction cast by the corresponding base classifier. In the “stringent” version we suppressed any instance of a mutation that had previously been used in the training split, whilst in the “permissive” version we allowed them.

#### Rare CH mutations from MSK-IMPACT

A set of rare mutations from the MSK-IMPACT dataset were obtained as a set for the validation analysis (10). This set of rare mutations consists of SNVs from the 12 CH genes with boostDM-CH models obtained by Gao *et al*., but not included in any of the discovery cohorts, and thus, absent from the training set. As a negative set for this validation, we used synthetic mutations simulated using the MSK-IMPACT cohort mutational profile. For each gene, we selected 50 sets including unique mutations with balanced labels (same number of positive and negative examples) and annotated them with the prediction of the corresponding boostDM-CH general model. Only genes with more than 5 unique mutations were included in the validation study.

#### Biobank Japan CH mutations

We obtained a dataset of mutations from the DNA Data Bank of Japan (DDJB) or Biobank Japan (29) through accession numbers JGAS000293/JGAD000399 and JGAS000293/JGAD000400. From this independent set of observed mutations, described in Saiki *et al*., 2021 (56) we extracted the SNVs from the 12 genes with boostdm-CH models. For the negative set, we used synthetic mutations generated in the three discovery cohorts. For each gene, we selected 50 sets including unique mutations with balanced labels (same number of positive and negative examples) and annotated them with the prediction of the corresponding boostDM-CH general model. Only genes with more than 5 unique mutations were included in the analysis.

### DNMT3A mutants methylation efficiency

We used Nicholas *et al*., 2023 (38) methylation activity data across *DNMT3A* mutants. They integrated base editing by single-guide RNA with a DNA methylation reporter to perform *in situ* mutational scanning of *DNMT3A* in cells to evaluate the impact of 156 unique missense/nonsense *DNMT3A* variants in the methylation activity. Variants with a sgRNA score >2 s.d. (within cells selected by their diminished level of methylation of a reporter gene) above or below the mean of intergenic negative controls were considered ‘enriched’ or ‘depleted,’ within the pool of all guides, respectively.

### Expert-curated rules to identify CH driver mutations

Three sets of expert-curated rules designed by CH researchers (referred to as Niroula (25); Bick (26), and WHO (27)) were used for boostDM-CH performance comparison. For such benchmarking we only consider those mutations in genes included by all the three sets of expert-curated rules (10 genes).

### Comparison with myeloid boostDM models

The myeloid boostDM models were trained with data from 11 tumor-type specific cohorts from IntOGen (Release v2023.05.31) (33) comprising Acute Myeloid Leukemias (9 cohorts), Myelodysplastic Syndromes (1 cohort) and Chronic Myelogenous Leukemias (1 cohort).

### Application to population-based cohort (UK Biobank)

All analyses were performed with the UK Biobank Research Analysis Platform (22). The cohort used in the study comprises 469,880 individuals for whom whole exome sequencing (WES) data was readily available. Individuals with hematological neoplasms at baseline (that is, with a hematological cancer diagnosis date before the date they attended the assessment centers) were excluded from the analysis, yielding 467,202 individuals included in the analysis (age range: 37–73, median age: 58 years old; 54% females).

### Identification of CH mutations across UKB donors

CRAM files generated by the OQFE pipeline from UKB were used. All the following steps were carried out within a nextflow (54) pipeline (v20.10.0). Variant calling on WES data from 467,202 individuals was performed using Mutect2, Genome Analysis Toolkit (GATK, v.4.2.2.0) (57). Briefly, Mutect2 was run in ‘tumor-only’ mode with default parameters, for a mini-BAM comprising the sequences of the exons of 11 with boostDM-CH models (excluding *U2AF1*, with a known artifact in the GRCh38 assembly of the human genome). A panel of normals was obtained from 100 WES from the youngest individuals (<41 years old at recruitment) in UKB, generating a blacklist of mutations present in at least 2 of these individuals. Raw variants called by Mutect2 were annotated with FilterMutectCalls using the estimated prior probability of a reading orientation artifact generated by LearnReadOrientationModel (GATK, v.4.2.2.0) (58), even though no filters were applied using this annotation. Since variant calling pipelines such as Mutect2 cannot reliably identify variants in *U2AF1* in sequencing data that are mapped to the human GRCh38 (hg38) reference genome due to an erroneous duplication of the *U2AF1* locus in this reference genome, a custom script was used to identify variants in *U2AF1*. First we performed a pileup approach to obtain all counts of mutated alleles present in reads that are mapped at any of the two U2AF1 genomic loci (ENST00000291552 chr21:43092956-43107570 and ENST00000610664 chr21:6484623-6499248) and then we annotated this mutated reads for a unique genomic location for ENST00000291552. Gene annotation was performed using Ensembl Variant Effect Predictor (VEP) (v.101) (52). To filter out potential germline variants we used a population reference of germline variants generated from the 1000 Genomes Project (1000GP, Phase 3) (59) and the Genome Aggregation Database (gnomAD, r2.1.1) (60). The identified variants were subjected to the following filters: only SNVs were selected, we required a minimum number of alternate reads of 3, a maximum of 2 altered alleles, and a MAF lower than 0.001 according to 1000GP (EUR_AF) or gnomAD (gnomAD_AF_NFE, gnomAD_AF). Additionally, for *U2AF1* mutations we selected those with evidence of the variant on both forward and reverse strands.

### Associations of CH with age and other phenotypes in the UKB

Association analyses of CH with several conditions and risk analyses were performed on the DNAnexus platform of the UKB. These analyses were performed taking into account different subsets of UKB donors (reflected in Figures): 1) carriers of one CH driver mutation according to boostDM-CH models; 2) carriers of more than one CH driver mutation (multiple drivers); 3) carriers of a CH driver mutation at VAF>=10% (driver large); 4) carriers of a CH driver mutation at VAF<10% (driver small); 5) carriers of non-driver mutations (non-driver); 6) carriers of any potential blood mutation (irrespective of its classification) detected in the somatic calling (potential mutation). For some analyses, we grouped carriers of CH driver mutations in genes of different functional groups, such as **chromatin modifiers** (*DNMT3A*, *TET2* and *ASXL1*), **DNA damage response** (*TP53*, *CHEK2*, *PPM1D* and *MDM4*) and **splicing factors** (*SF3B1*, *SRSF2* and *U2AF1*).

#### Phenotypes

Clinical data from the UKB was downloaded in November 2022 and individual traits were pulled out from the whole phenotype file classified in data-fields. Basic information from the individuals used for the analyses was age of recruitment (data-field: 21022), sex (data-field: 31), genetic principal components (data-field: 22009), death status (data-field 40007), and body mass index (BMI, data-field: 21001). Smoking status was defined as never smoker or ever smoker using smoking status information (data-field: 20116).

Presence of cancer was defined from reported occurrences of cancer (data-field: 40009). Years to first cancer was assessed using the age of recruitment and the age of first cancer (data-field: 22008). Specific cancer type, cardiovascular diseases (CVD) traits, infectious disease and other conditions such as hypertension or diabetes mellitus type II were generated combining information from different data-fields (Table S2) including ICD-10 diagnosis (data-fields: 40006, 41202, 41270), ICD-9 diagnosis (data-fields: 40013, 41203, 41271), self-reported cancer (data-field: 20001), self-reported non-cancer illness (data-field: 20002), underlying cause of death (data-field: 40001), contributory cause of death (data-field: 40002), operation (data-field: 20004), and OPSC4 (data-field: 41272, 41200), similarly definitions outlined by Siddhartha *et al*. 2020 (61) and Trinder *et al.* 2020 (62). For each definition, the first diagnosis event that occurred was selected. Years to first occurrence of cancer, CVD and infectious diseases was calculated also using the difference between date of recruitment and specific diagnosis dates (data-fields: 40005, 40000, 41260, 41262, 41263, 41280, 41281, 41282) and diagnosis age (data-fields: 20007, 20009) based on disease definitions. Regarding some of the covariates used for the association, diabetes mellitus type II was defined as its diagnosis or treatment with insulin or oral hypoglycemic medication (data-field: 6177); dyslipidemia was defined as cholesterol ≥240 mg/dL (data-field: 30690), LDL-direct ≥160 mg/dL (data-field: 30780), HDL-cholesterol <40 mg/dL (data-field: 30760), or use of lipid-lowering drugs (data-field: 6177); hypertension was defined by its diagnosis or by having a systolic blood pressure ≥140 mmHg (data-field: 4080), diastolic blood pressure ≥90 mmHg (data-field: 4079), or use of antihypertensive medication (data-field: 6177).

#### Analyses

Unless otherwise specified, all regression models included age, sex, and the first ten ancestry principal components as covariates. For cancer regressions, we also included smoking status as covariate, while for CVD we included smoking status, BMI, diabetes mellitus type II, dyslipidemia, and hypertension status. Infectious diseases included smoking status and hematological cancer. For age, cancer, smoking status, CVD, and death association with CH we performed logistic regression analysis using logit (Python statsmodel package v.0.14.0). To analyze the risk of having hematological malignancies and myeloid neoplasms in CH carriers, we performed a Cox proportional hazards model CoxPHFitter (Python lifelines package v.0.27.8). We count as an event any reported diagnosis of hematological or myeloid cancer, respectively, after enrollment in the UKB. Individuals without the event who died before the end of the follow-up were censored at the time of death, while the rest were censored at the last follow-up reported (2021-06-25, from data-field 40005). The maximum number of years to an event was restricted to the 97th percentile of the UKB population. Kaplan-Meier curves were performed using KaplanMeierFitter and logrank_test functions (Python lifelines package v.0.27.8).

### Estimation of CH mutation fitness

Using a quantitative framework, Watson *et al.* 2020 (44) applied population genetic theory to estimate the fitness advantage conferred by specific mutations in blood by analyzing the spectrum of VAFs from sequencing data. Using the same approach, we aimed to infer the fitness score of the most frequent SNVs from the UKB dataset (>=30 observed mutations). Fitness estimation was performed using a custom Python script based on the approach developed by Watson *et al*., 2020. Briefly, probability density histograms, as a function of log VAFs, were plotted using Doane’s method for log VAF bin size calculation and then applying a maximum likelihood approach, fitness (s) was inferred. Data was trimmed applying gene-specific VAF detection threshold below which the density began to decline. From the initial 1028 recurrent SNVs, we exclude those with a multimodal VAF distribution (p-value > 0.2), median VAF >0.2 or with VAF values above the limit of detection set (see above) that resembled a normal distribution (p-value < 0.05) in order to exclude possible germline variants. We were thus able to compute fitness scores for 220 SNVs (136 drivers and 84 non-driver CH mutations, according to boostDM-CH).

### Data and software availability

Blood somatic mutation data required to train boostDM-CH models is available through Hartwig Medical Foundation (HMF) and dbGaP following the same procedure to access the original datasets used in the reverse calling approach. HMF blood somatic mutations are available as part of the data access request to HMF (https://www.hartwigmedicalfoundation.nl). TCGA blood somatic mutations are available through dbGaP (phs002867) to researchers who have obtained permission to access protected TCGA data. Panel-sequenced data from the IMPACT targeted cohort is available through cBioPortal (https://www.cbioportal.org/study/summary?id=msk_ch_2020). The compendium of CH driver genes is available through IntOGen (intogen.org/ch), as is the *in silico* saturation mutagenesis of CH drivers (intogen.org/ch/boostdm). Data required to reproduce UKB and Japanese Biobank analyses is available upon access request to both entities (details above). Software required to train boostDM-CH models and carry out analyses described in the article will be publicly available upon publication.

## Data Availability

All data produced in this study is available at www.intogen.org/ch/boostdm

## Acknowledgements

The authors want to acknowledge Martina Gasull and Paula Gomis for support in data access and Federica Brando for key support in the development of the boostDM-CH website. N.L.-B. acknowledges funding from the European Research Council (consolidator grant 682398). S. D. was supported by a Juan de la Cierva fellowship from Spanish Ministerio de Ciencia e Innovación (IJC2020-044728-I). J. E. R-Z. was supported by a Postdoctoral AECC 2023 fellowship from fundación científica asociación Española Contra el Cáncer (AECC) (POSTD234814RAMI). This project was supported by the CHEMOHEALTH project, funded by the Spanish Ministry of Science (MCIN), AEI/10.13039/501100011033/ and by FEDER, the AtheroClonal project, also funded by the Spanish Ministry of Science (MCIN), AEI and the European Union “NextGenerationEU”/PRTR”, and the MyoClonal project, funded by la Caixa, HR22-00732. It has also been supported by the project “Discovering the molecular signatures of cancer PROMotion to INform prevENTion” (PROMINENT) funded by Cancer Research UK (CGCATF-2021/100008), National Cancer Institute (1OT2CA278668-01) and the Spanish Cancer Association, AECC. IRB Barcelona is a recipient of a Severo Ochoa Centre of Excellence Award from the Spanish Ministry of Economy and Competitiveness (MINECO; Government of Spain) and an Excellence Institutional grant by the Asociacion Española contra el Cancer, and is supported by CERCA (Generalitat de Catalunya). This publication and the underlying research are partly facilitated by Hartwig Medical Foundation and the Center for Personalized Cancer Treatment (CPCT) which have generated, analyzed and made available data for this research. This publication and the underlying research are also partly facilitated by data collected and made public by The Cancer Genome Atlas network. This research has been conducted using the UK Biobank Resource under Application Number 69794.

## Author contributions

SD and JER-Z, carried out most analyses of the results of the application of the boostDM-CH models to the mutations in the discovery cohorts, validation cohorts and UKB. FM carried out the training and validation of boostDM-CH models and contributed to downstream analyses. SD, JER-Z, and FM generated the figures in the manuscript. MLG carried out the identification of somatic mutations in 12 CH driver genes across UKB donors. MA carried out the analysis of fitness of recurrent CH mutations across UKB donors. SD, FM, AG-P, and NL-B conceived the project. AG-P and NL-B supervised the project and its analyses and prepared the first draft of the manuscript. All authors contributed to the discussion of results and the completion of the manuscript.

## Authors’ disclosures

The authors declare no potential conflicts of interest.

## Notes

### Competing Interest Statement

The authors have declared no competing interest.

### Author Declarations

Blood somatic mutation data required to train boostDM-CH models is available through Hartwig Medical Foundation (HMF) and dbGaP following the same procedure to access the original datasets used in the reverse calling approach. HMF blood somatic mutations are available as part of the data access request to HMF (https://www.hartwigmedicalfoundation.nl). TCGA blood somatic mutations are available through dbGaP (phs002867) to researchers who have obtained permission to access protected TCGA data. Panel-sequenced data from the IMPACT targeted cohort is available through cBioPortal (https://www.cbioportal.org/study/summary?id=msk_ch_2020g). Data in the UK Biobank and Japanese Biobank analyses is available upon access request to both entities (https://www.ukbiobank.ac.uk/enable-your-research/apply-for-access and https://biobankjp.org/en/info/offer.html, respectively)

