## Supplemental Material for "Identification of Clonal Hematopoiesis Driver Mutations through In Silico Saturation Mutagenesis"

### Supplementary Information

### Table of contents

**Supplementary Methods** **3**
BoostDM-CH models 3
Discovery index 3
Construction of positive and negative sets of mutations 4
Features of mutations in CH genes 5
Training boostDM-CH models 6
**Supplementary Note** **9**
**Supplementary Figures** **10**
**Supplementary Tables** **45**
**References** **53**

### Supplementary Methods

#### BoostDM-CH models

BoostDM-CH models are ensembles of base classifiers (n=50) each trained with a random sample subset drawn from the learning data. Each base classifier is a boosted trees model (trained with XGBoost gradient boosting implementation) (1) with a cross-entropy loss function. The base classifiers are aggregated into a consensus model with an aggregator intended to correct for the systematic bias of each expert classifier. Each base classifier also produced SHAP local explanations (see main Methods and (2)) and the consensus model summarizes them with the mean.

#### Discovery index

We were interested in estimating the expected count of unique mutations as a function of the number of sequenced samples, which we can denote by E(n). This gives us a way to numerically represent the extent to which some driver mutations may have not yet been discovered, thereby depicting the discovery status of mutations per gene.

We define a continuous score, the discovery index, to track the probability that, as new samples are sequenced, mutations that affect the gene have been identified previously. This score takes values in the unit interval, where higher values represent that fewer previously unobserved mutations are expected if new samples are to be sequenced. The discovery index aims to encode the bend of the function E(n), which in turn indicates how far/close from having seen all the possible driver mutations we are at the moment.

To this end we generated a collection of data points (n,u) by drawing random subsets with size n and counting the number of unique mutations observed as u. For each gene we proceeded by repeatedly sampling subsets of samples and counting unique mutations. To do so, we scanned a grid of 20 different sample sizes, evenly spaced between zero and the total number of samples for which we have sequenced the gene. For each size n, several samples (k = 100) were drawn independently with replacement.

To compute the discovery index, we repeatedly (100 times) bags of data points comprising one point per each number of samples, then derived the best (weighted) least-squares fitting curve of the form:

$$f(n) = \alpha(1 - \exp(\beta n))$$

We define the discovery index of a single fit as:

$$d_i = \min\{f_i(N)/\hat{\alpha}_i, 1\}$$

with N being the total number of samples sequenced for the gene. We choose a weighting to conduct the fitting that favors points (n) where the variance of the distribution of u's is higher:

$$L(n, f(n)) = \sum_{n \in Grid} w_n (n - f(n))^2, \text{ with } w_n \propto 1/(1 + \sigma(n)^e)$$

where $\sigma(n)$ is the standard deviation of the u-values at n. Finally, we computed the discovery index at each gene as the median across the 100 different resampling and fitting steps.

#### Construction of positive and negative sets of mutations

*Excess of driver mutations*

We computed the so-called excess of mutations for a given coding consequence-type as the proportion of observed mutations matching this consequence-type that are not explained by the neutral mutation rate. The excess is inferred from the consequence-type specific dN/dS estimate $\omega$ as $e = (\omega - 1)/\omega$. We computed the excess for missense, nonsense and splicing-affecting mutations.

*Training set*

The first requirement for our supervised learning approach is to create a catalog of mutations labeled as either "likely drivers" or "likely non-drivers". With the table of mutations as vectors of features and binary driver response variable, we proceed to split the data and generate distinct base models from each split.

*Positive set*

We deem likely driver mutations those observed mutations with high chances of being driver mutations. We based this decision on the consequence-type specific excess derived from dNdScv (3). If a mutation has a consequence type whose excess of observed-over-expected mutations is higher than 80% we label the mutation as a likely driver and use it as a positive instance in the training set. Repeated mutations (i.e., when the same mutation is observed in different samples) are allowed in the positive set.

*Negative set*

For each likely driver mutation mapping to a gene and cohort of samples, we randomly select one mutation from the CDS (VEP.92 canonical transcript) (4) with probabilities dictated by the trinucleotide context of the mutation and the background mutational profile of the cohort. Because we split the data 50 times, we build the negative set by performing such a random selection 50 times with replacement.

*Training splits*

Each base classifier is obtained after conducting training with two sets of annotated mutations: "train" and "test". We will refer to a train-test pair as a split. In our setting each split is generated randomly and must satisfy the following requirements:

- Both train and test sets have to be balanced (same positive and negative items).
- The train set comprises 70% of the total positive set (allowing repeated mutations).
- Each unique positive instance belongs either to train or test, but not both.
- Repeated positive mutations are allowed in train, but not in test, to prevent spurious inflation of the cross-validation performance.

- Non-driver mutations in train and test are randomly selected among all the mutations in the pool of non-driver mutations.

We set a minimum average number of mutation instances in the train set at 30. Otherwise the model is not computed.

### Features of mutations in CH genes

Given a point mutation in a driver gene and tumor type (either observed or randomized) our method requires an annotation of the mutation with a series of features to train the classifiers.

*Consequence type*
For every mutation its consequence type in the canonical transcript was extracted from VEP v.92 as a Sequence Ontology (SO) term. Whenever a mutation was associated with several consequence types, we kept the most severe according to the order of severity defined by Ensembl: [https://www.ensembl.org/info/genome/variation/prediction/predicted_data.html](https://www.ensembl.org/info/genome/variation/prediction/predicted_data.html). We annotated each mutation with a categorical variable with 4 levels: “missense” (SO: missense_variant), “nonsense” (SO: stop_gained), “splicing” (SO: splice_donor_variant, splice_acceptor_variant, splice_region_variant) and “synonymous” (SO: synonymous_variant). We encoded this as 4 dummy variable features: csqn_type_synonymous, csqn_type_missense, csqn_type_nonsense, csqn_type_splicing.

*Clustering features*
Four features were derived from the statistical analysis of blood somatic mutations across more than 12,000 donors from large cancer genomics cohorts and more than 24,000 targeted sequenced samples as part of Intogen-CH (5,6): two features reflecting linear clusters in the DNA primary structure, one feature reflecting clusters of mutations on the protein 3D structure and another feature reflecting the enrichment of mutations in Pfam domains.

CLUSTL_cat_1: We define a dichotomous feature reflecting whether a non-synonymous mutation overlaps a significant linear cluster identified by the method OncodriveCLUSTL (7) in any of the three somatic blood mutation cohorts.

CLUSTL_SCORE: Additionally we create another feature that represents the maximum OncodriveCLUSTL score achieved across linear clusters overlapping the mutation.

HotMaps_cat_1: We define a dichotomous feature reflecting whether a missense mutation overlaps a significant cluster of mutations on the protein 3D identified by the method HotMAPS (8) using a custom implementation that takes the neutral mutational profile into account.

smRegions_cat_1: We define a dichotomous feature reflecting whether a non-synonymous mutation overlaps a Pfam domain (9) that is significantly enriched for mutations in the gene identified by the method smRegions (10).

*Phylogenetic conservation*
Each mutation was annotated with the conservation of the reference nucleotide across vertebrates measured through the PhyloP 100-way score (11).

*Post-translational modifications*
We define 5 dichotomous features reflecting whether non-synonymous mutations were annotated as being subject to post translational modifications (PTMs) of each of the following types according to the PhosphositePlus database (12): acetylation, phosphorylation, ubiquitination, methylation or any other regulatory modification.

*Nonsense-mediated RNA decay*
We define a dichotomous feature reflecting whether a nonsense mutation overlaps the first or last coding exon of the canonical transcript (according to VEP.92) reflecting the potential for the truncating variant to skip nonsense-mediated RNA decay (13).

### Training boostDM-CH models

Gradient boosting learning with tree functions (boosted trees) aims to find the tree function T (sum of trees) that minimizes the empirical expected loss between the observed and predicted response values (risk minimization):

$$\hat{T} = argmin_{T} \frac{1}{N} \sum_{i=1}^{N} L(y_i, T(x_i))$$

where x, y are the feature vectors and binary responses, respectively, and $L(y, \hat{y})$ is a custom loss function measuring the gap between the true label and the predicted probability.

**Loss**
In our context of classification we choose as custom loss function is the negative log-likelihood of the Bernoulli distribution, also known as log-loss or cross-entropy:

$$L(y, \hat{y}) = - y \cdot log(\hat{y}) - (1 - y) \cdot log(1 - \hat{y}).$$

This is encoded in the XGBoost hyperparameter configuration as:

```
"objective": "binary:logistic"
```

**XGBoost config**
The model hyperparameters specify the learning strategy to keep a balance between loss minimization and generalization. While some hyperparameter values are the result of an explicit design decision, others cannot be unequivocally defined. The current values stand as a compromise resulting from a testing series. For more information about hyperparameter tuning with XGBoost, please check xgboost documentation: xgboost.readthedocs.io.

```
XGB_PARAMS = {
```

"objective": "binary:logistic",
"reg_lambda": 1,
"random_state": 42,
"scale_pos_weight": 1,
"subsample": 0.7,
"reg_alpha": 0,
"max_delta_step": 0,
"min_child_weight": 1,
"learning_rate": 1e-03,
"colsample_bylevel": 1.0,
"gamma": 0,
"colsample_bytree": 1.0,
"booster": "gbtree",
"max_depth": 4,
"silent": True,
"seed": 21
}

See below a more detailed explanation of the choice of different training parameters.

**Early stopping**

For training each base classifier (coming from each split) we set a maximum of 20,000 estimators for each base model. To prevent overfitting, we established an early-stopping criterion so that the training stops whenever the performance in the test set is not improving (log-loss does not decrease) for 2,000 iterations.

**Cross-validation**

For each base classifier we computed the cross validation performance as a weighted version of the F-score (harmonic mean between precision and recall) that gives precedence to precision over recall, which we refer to as $F_{50}$. If we denote the precision and recall as P and R, respectively, the $F_{50}$ is computed as:

$$F_{50} = 1.25 \cdot PR/(0.25 \cdot P + R)$$

**Forecast aggregator**

The final predictions of a model is computed as consensus of the prediction of the 50 base classifiers. Our approach resorts to a non-linear combination of probabilities based on a logit-normal model (14,15). Specifically, if the i-th base classifier casts a prediction $p_i$ that a specific mutation is a driver and we set $Y_i = logit(p_i)$, our modeling choice assumes that the prediction $p_i$ arises from some latent true probability $p$ that the mutation is a driver, which in turn has been perturbed by some degree of systematic bias:

$$Y_i = \log\,(p/(1-p))^{1/a} + \epsilon_i, \text{ with } \epsilon_i \sim N(0, \sigma^2)$$

where $a \geq 1$ represents the amount of systematic bias.

Intuitively, the systematic bias quantifies the extent to which individual classifiers regress towards a log-odds zero due to partial information or under-confidence while issuing their respective forecasts. While $a = 1$ would be associated with an accurate forecast, $a > 1$ would represent under-confidence.

Using the stated modeling assumptions, given a collection of classifiers and their respective forecasts $p_i$ and logits $Y_i$, the latent true probability $p$ can be computed with the following estimator:

$$\hat{p} = \exp(a\overline{Y})/(1 + \exp(a\overline{Y}))$$

where $\overline{Y}$ denotes the average of the logits and $a$ is the amount of systematic bias. The systematic bias gives us a way to sharpen the consensus forecasts of a coalition of under-confident predictors. In the interest of clarity and interpretability of the outcome, we require boostDM forecasts to have a marked bimodal forecasting propensity, yielding forecasts close to either 0 or 1. We run boostDM with $a = 2.3$ as systematic bias.

### Supplementary Note

#### Considerations on the use of UKB mutations for the validation of boostDB-CH models

While we use the identification of CH driver mutations in the UKB and their association with age or the risk to develop several phenotypes as a validation of the performance of boostDM-CH models, we reasoned that recurrent CH driver mutations identified in this cohort probably shows a high degree of overlap with the set of blood somatic mutations in the discovery cohorts. Since these are the mutations used to train boostDM-CH models, we hypothesized that a certain bias in this validation attributable to this overlap could occur. Recurrently observed CH mutations are more likely to be drivers (providing higher fitness and stronger association with age). Their identification as such by boostDM-CH models therefore does not imply overfitting, but it does beg the question of how much they are generalizing the identification of driver mutations beyond the realm of the training set.

To investigate further the performance of boostDM-CH models in UKB, we decided to focus on the most rare CH driver mutations, which are less likely to be classified by the models just because they appear in the discovery cohorts used to train them. Specifically, 16,908 (41%) CH driver mutations in UKB had never been observed in any of the three discovery cohorts, while 4,975 (9%) had been observed only once (Fig. S17a). These non-observed or non-recurrent mutations are significantly associated with age (Fig. S17b) and the estimated fitness of the subset that are recurrent in UKB is significantly greater than that estimated for recurrent non-driver mutations (Fig. S17c). This shows that the models are indeed capable of identifying new CH driver mutations beyond highly recurrent ones, which are likely to be part of their training.

These two findings constitute signs that these mutations are indeed novel (or very rare) CH drivers identified by boostDM. This, in turn, (and in addition to the validations carried out in the sets of mutations in Biobank Japan and the work by Gao *et al*., 2021 (16)) provides evidence that the models are capable of generalizing in the task to identify these mutations rather than memorizing the training set.

### Supplementary Figures

Figure S1

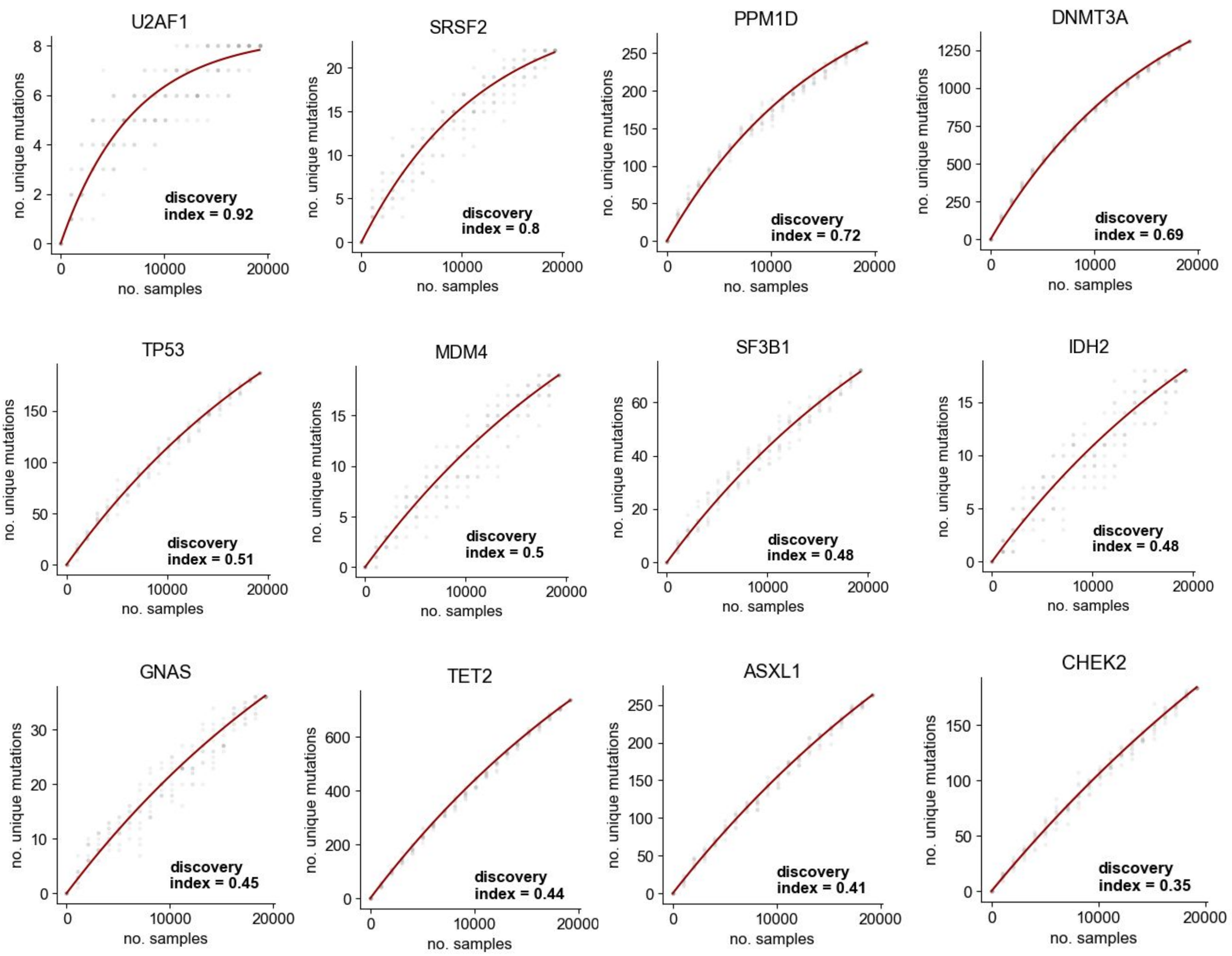
U2AF1
discovery index = 0.92
SRSF2
discovery index = 0.8
PPM1D
discovery index = 0.72
DNMT3A
discovery index = 0.69
TP53
discovery index = 0.51
MDM4
discovery index = 0.5
SF3B1
discovery index = 0.48
IDH2
discovery index = 0.48
GNAS
discovery index = 0.45
TET2
discovery index = 0.44
ASXL1
discovery index = 0.41
CHEK2
discovery index = 0.35
no. unique mutations
no. samples

**Figure S1. Discovery index of the 12 high-quality boostDM-CH gene-models.** To compute the discovery index, subsets of samples of the discovery cohorts are randomly chosen and the number of unique mutations of each gene in each subsample is counted. The trend resulting from these random subsamples (dots) are then computed (red lines) and the discovery index is subsequently computed (Methods).

### Figure S2

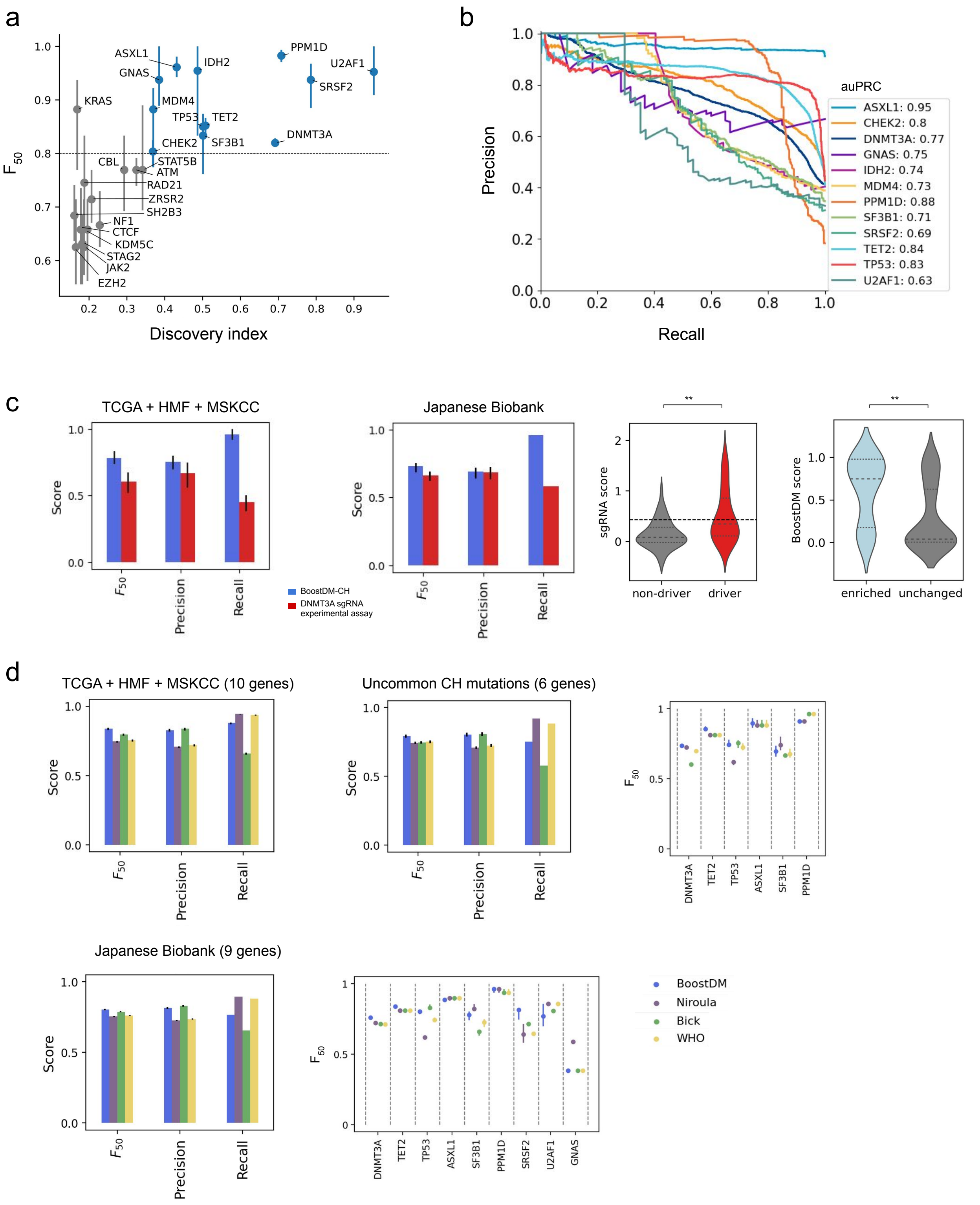

a
ASXL1
GNAS
IDH2
PPM1D
U2AF1
SRSF2
KRAS
MDM4
TP53
TET2
DNMT3A
CHEK2
SF3B1
CBL
STAT5B
ATM
RAD21
ZRSR2
SH2B3
NF1
CTCF
KDM5C
STAG2
JAK2
EZH2
Discovery index
b
Precision
Recall
auPRC
ASXL1: 0.95
CHEK2: 0.8
DNMT3A: 0.77
GNAS: 0.75
IDH2: 0.74
MDM4: 0.73
PPM1D: 0.88
SF3B1: 0.71
SRSF2: 0.69
TET2: 0.84
TP53: 0.83
U2AF1: 0.63
c
TCGA + HMF + MSKCC
Japanese Biobank
Score
Precision
Recall
BoostDM-CH
DNMT3A sgRNA experimental assay
sgRNA score
non-driver
driver
BoostDM score
enriched
unchanged
d
TCGA + HMF + MSKCC (10 genes)
Uncommon CH mutations (6 genes)
Japanese Biobank (9 genes)
BoostDM
Niroula
Bick
WHO
DNMT3A
TET2
TP53
ASXL1
SF3B1
PPM1D
SRSF2
U2AF1
GNAS

**Figure S2. Performance of boostDM-CH models.**

a) Performance (median ± interquartile range (IQR) $F_{50}$) of the cross-validation (of 50 base classifiers) of the 25 initial CH models; similar as Figure 1d, but as a function of the discovery index computed for the driver gene.

b) Area under the Precision-Recall curves of the 12 high-quality boostDM models; similar to Figure 1e, but using unique mutations in the cross-validation splits (see Methods).

c) Comparison of the performance (several metrics) of boostDM-CH DNMT3A model and an experimental DNMT3A base edition assay (17) on mutations of the three discovery cohorts (first bar plot, corresponding to the bars shown in Fig. 1f) and another independent set of donors (second bar plot). The first violin plot presents the distribution of sgRNA scores (estimating the decrease in DNMT3A methylation activity of mutants generated through a base edition assay) of driver and non-driver DNMT3A mutations according to the boostDM-CH model. The second violin plot presents the distribution of boostDM-CH scores of DNMT3A mutants found to be significantly (enriched) or non significantly less active (unchanged) in a methylation reporter assay than an intergenic mutant used as negative control.

d) Left, distribution of methylation scores (per the aforementioned base edition assay) of boostDM drivers and non-driver mutations. Right, distribution of boostDM-CH scores of mutations that change (enriched) or not the methylation capability of DNMT3A.

e) Performance of the classification of blood somatic mutations of boostDM-CH models and three expert-curated rules (18–20) across three different CH datasets. Barplots represent overall performance (across 10, 9 or 6 genes, depending on the dataset), while the dot plots depict the performance of each gene-specific model. In each bar plot, the first set of bars ($F_{50}$) corresponds to those shown in Figure 1g.

### Figure S3

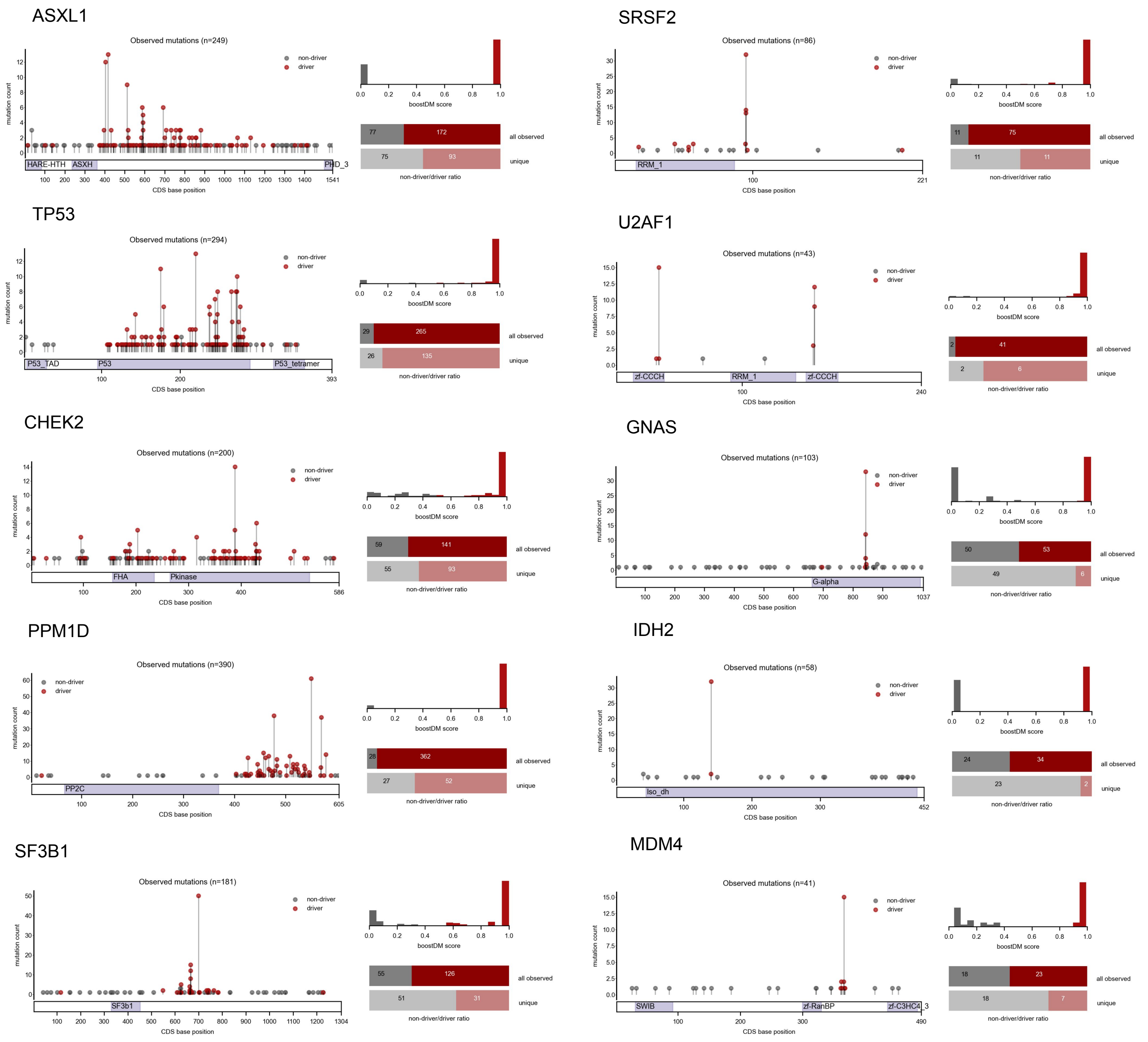

ASXL1
Observed mutations (n=249)
non-driver
driver
mutation count
HARE-HTH
ASXH
PHD_3
CDS base position
boostDM score
77
172
all observed
75
93
unique
non-driver/driver ratio
SRSF2
Observed mutations (n=86)
RRM_1
11
75
11
11
TP53
Observed mutations (n=294)
P53_TAD
P53
P53_tetramer
29
265
26
135
U2AF1
Observed mutations (n=43)
zf-CCCH
RRM_1
zf-CCCH
2
41
2
6
CHEK2
Observed mutations (n=200)
FHA
Pkinase
59
141
55
93
GNAS
Observed mutations (n=103)
G-alpha
50
53
49
6
PPM1D
Observed mutations (n=390)
PP2C
28
362
27
52
IDH2
Observed mutations (n=58)
Iso_dh
24
34
23
2
SF3B1
Observed mutations (n=181)
SF3b1
55
126
51
31
MDM4
Observed mutations (n=41)
SWIB
zf-RanBP
zf-C3HC4_3
18
23
18
7

**Figure S3. Distribution and classification of observed mutations along the protein sequence of 10 CH driver genes.**

Distribution of mutations observed in 10 CH driver genes (excluding DNMT3A and TET2, shown in Fig. 2a,b) in the three discovery cohorts along the sequence of each gene, with the color denoting their classification as driver or non-driver by boostDM-CH. The height of the needle representing each mutation corresponds to their recurrence across the three cohorts. Right-top, distribution of boostDM-CH scores of these observed mutations; right-bottom, proportion of all (dark colors) or unique (light colors) drivers and non-drivers out of all observed in each gene. Analogous to the top-right panels of Figure 2a and b.

### Figure S4

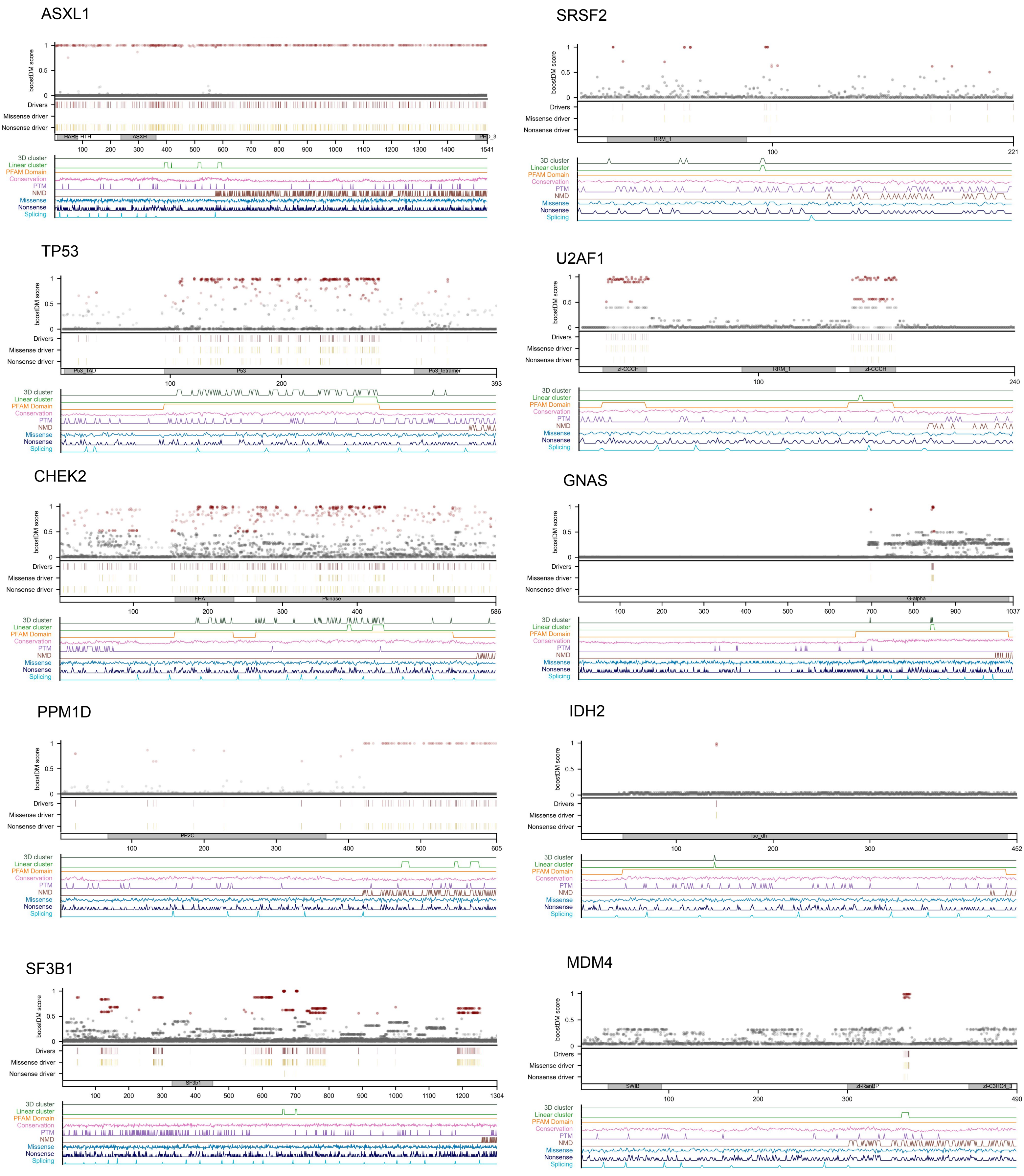
ASXL1
SRSF2
TP53
U2AF1
CHEK2
GNAS
PPM1D
IDH2
SF3B1
MDM4
boostDM score
Drivers
Missense driver
Nonsense driver
3D cluster
Linear cluster
PFAM Domain
Conservation
PTM
NMD
Missense
Nonsense
Splicing
HARE-HTH
ASXH
PHD_3
RRM_1
P53_TAD
P53
P53_tetramer
zf-CCCH
FHA
Pkinase
G-alpha
PP2C
Iso_dh
SF3b1
SWIB
zf-RanBP
zf-C3HC4_3

**Figure S4. In Silico Saturation Mutagenesis of 10 CH genes.**

The plots represent the distribution of driver and non-driver mutations among all possible mutations (*in silico* saturation mutagenesis) , with red representing predicted driver mutations (boostDM score >= 0.5) and gray, non-driver mutations. Ten genes are represented, excluding *DNMT3A* and *TET2*, already shown in Figure 2a and b. From top to bottom, the first plot presents boostDM scores of all possible SNVs along the protein sequence of each gene. The density of all potential SNVs classified as drivers is presented immediately below, with a distinction between missense and nonsense drivers (gold). The values of the mutational features used to train the models are shown linearly along the protein sequence in the plot at the bottom of the figure.

### Figure S5

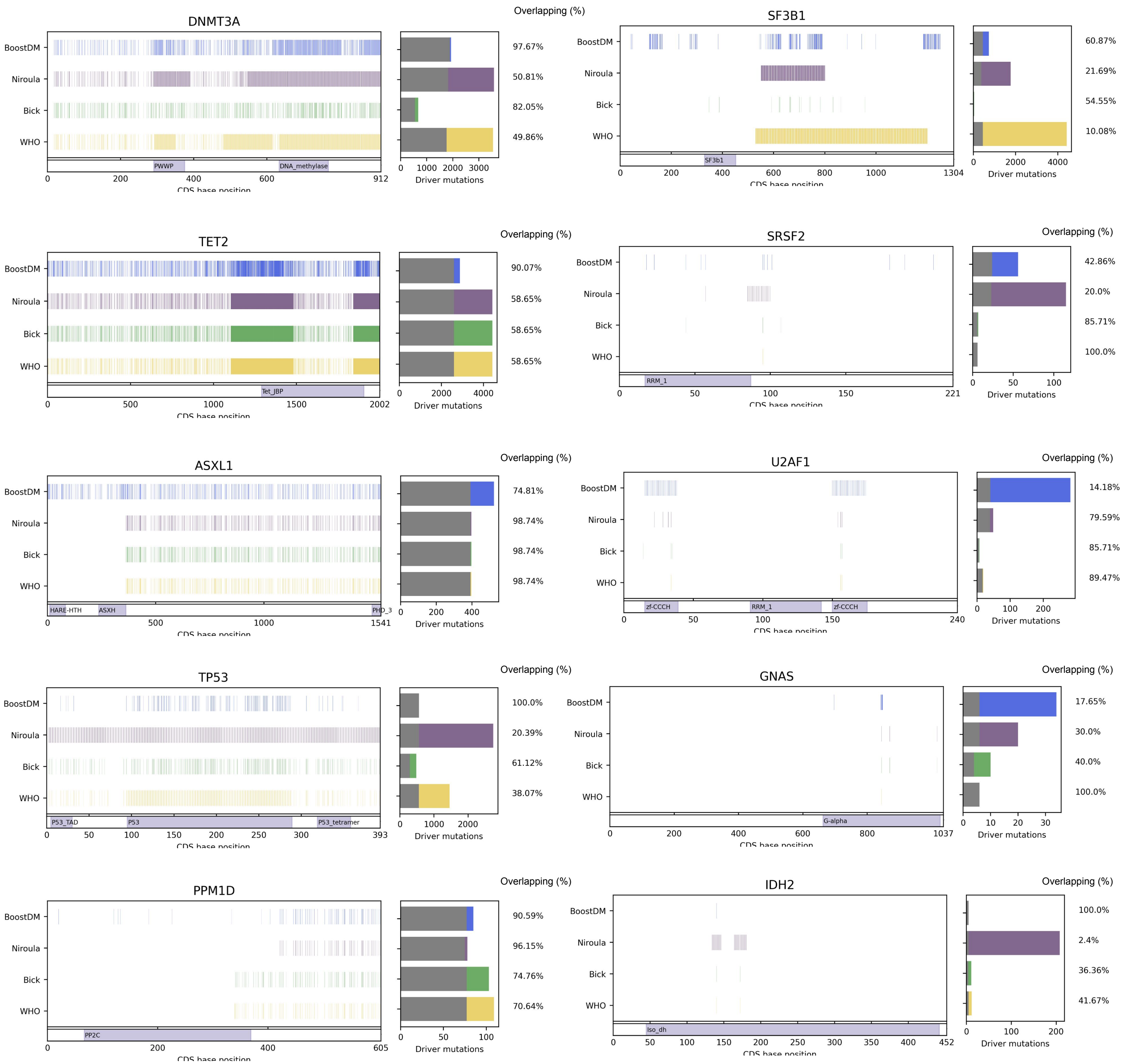
DNMT3A
BoostDM
Niroula
Bick
WHO
PWWP
DNA_methylase
CDS base position
Driver mutations
Overlapping (%)
97.67%
50.81%
82.05%
49.86%
SF3B1
SF3b1
60.87%
21.69%
54.55%
10.08%
TET2
Tet_JBP
90.07%
58.65%
58.65%
58.65%
SRSF2
RRM_1
42.86%
20.0%
85.71%
100.0%
ASXL1
HARE-HTH
ASXH
PHD_3
74.81%
98.74%
98.74%
98.74%
U2AF1
zf-CCCH
RRM_1
zf-CCCH
14.18%
79.59%
85.71%
89.47%
TP53
P53_TAD
P53
P53_tetramer
100.0%
20.39%
61.12%
38.07%
GNAS
G-alpha
17.65%
30.0%
40.0%
100.0%
PPM1D
PP2C
90.59%
96.15%
74.76%
70.64%
IDH2
Iso_dh
100.0%
2.4%
36.36%
41.67%

**Figure S5. Comparison of boostDM-CH models to prior expert-curated rules.**
Left boxes, distribution of driver mutations, among all possible SNVs in each gene, according to boostDM-CH (blue) or expert-curated rules (color code as in Figure 1) along the protein body (x-axis). Right boxes, proportion of mutations classified as drivers by boostDM-CH that are analogously classified as drivers by any of the prior expert-curated rule sets (top bar), or of mutations classified as drivers by one expert-curated rule set and boostDM-CH (other bars). In every bar, the gray segment represents the overlapping fraction, and the colored segments represent mutations uniquely classified by boostDM or a particular rule set.

Figure S6

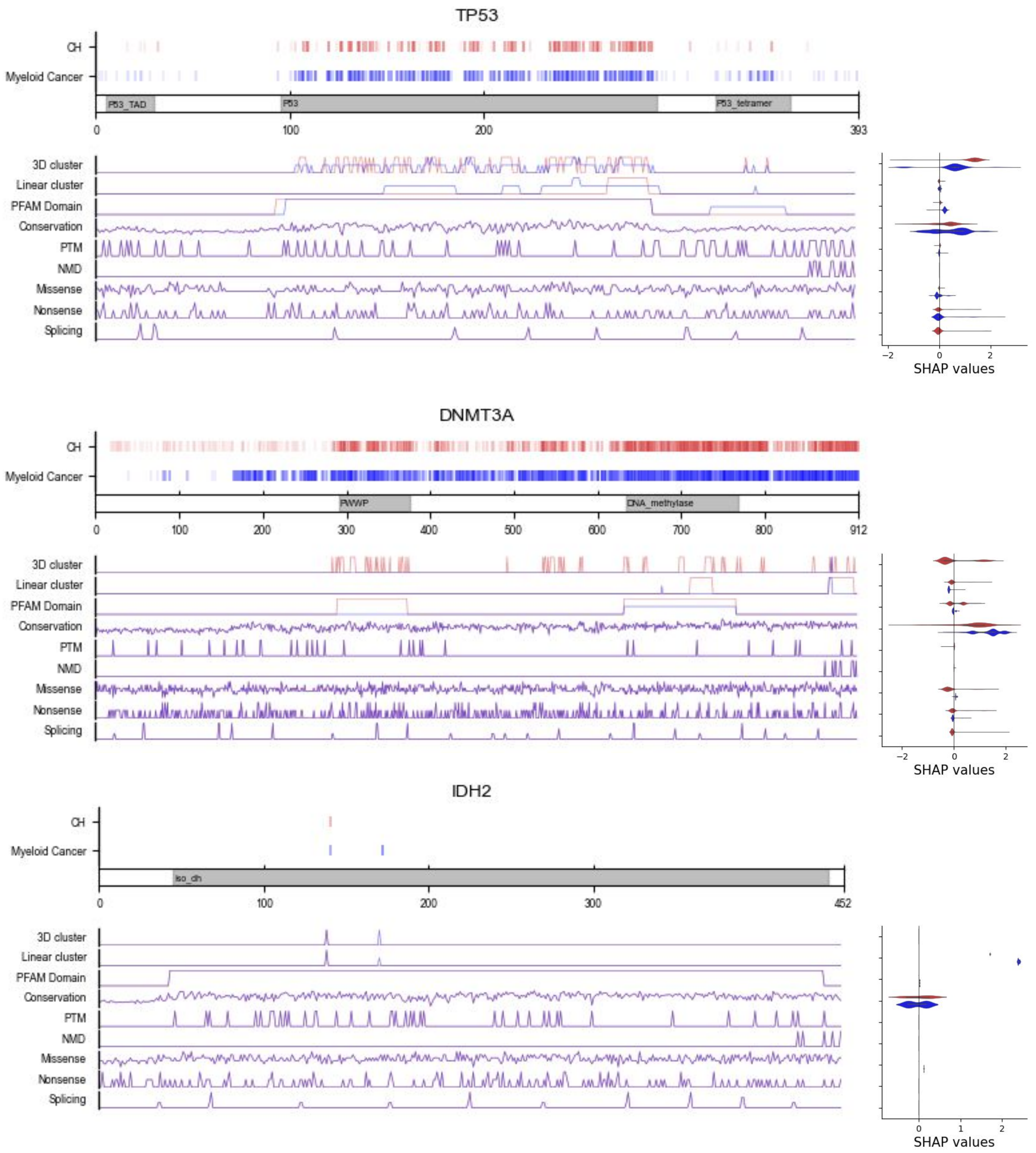
TP53
CH
Myeloid Cancer
P53_TAD
P53
P53_tetramer
0
100
200
393
3D cluster
Linear cluster
PFAM Domain
Conservation
PTM
NMD
Missense
Nonsense
Splicing
−2
0
2
SHAP values
DNMT3A
CH
Myeloid Cancer
PWWP
DNA_methylase
0
100
200
300
400
500
600
700
800
912
3D cluster
Linear cluster
PFAM Domain
Conservation
PTM
NMD
Missense
Nonsense
Splicing
−2
0
2
SHAP values
IDH2
CH
Myeloid Cancer
iso_dh
0
100
200
300
452
3D cluster
Linear cluster
PFAM Domain
Conservation
PTM
NMD
Missense
Nonsense
Splicing
0
1
2
SHAP values

**Figure S6. Comparison of boostDM-CH models and myeloid boostDM models of three CH driver genes.**

The top plot for each gene shows the distribution of driver mutations, among all possible mutations (*in silico* saturation mutagenesis), according to the boostDM-CH (red) or myeloid boostDM (blue) model of the gene. The values of mutational features used to train both types of models are shown linearly along the protein sequence in the plot at the bottom of the figure. The tracks are colored red if they represent the values of the features in boostDM-CH models, blue if they represent the values of features in myeloid boostDM models, and purple if they represent the overlap of the values of the features in both models. The violin plots at the right present the distribution of SHAP values of all driver mutations across the features used to train both types of models. Within the three genes (with the expectation of one *IDH2* recurrently mutated position, and several positions of *TP53*), the values of the features and the resulting predictions by the two types of models are very similar.

### Figure S7

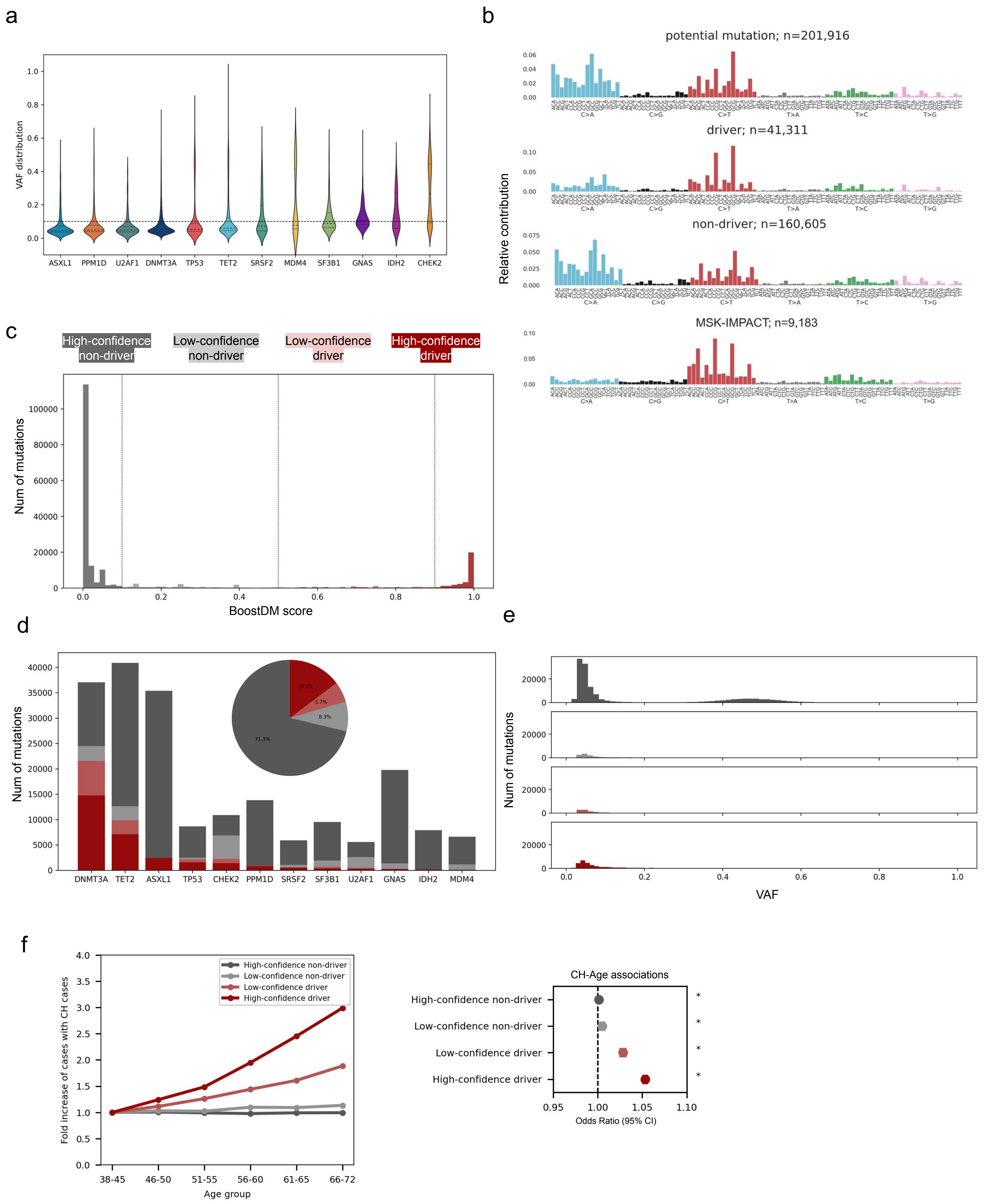
a
b
c
d
e
f
VAF distribution
ASXL1
PPM1D
U2AF1
DNMT3A
TP53
TET2
SRSF2
MDM4
SF3B1
GNAS
IDH2
CHEK2
potential mutation; n=201,916
driver; n=41,311
non-driver; n=160,605
MSK-IMPACT; n=9,183
Relative contribution
C>A
C>G
C>T
T>A
T>C
T>G
High-confidence non-driver
Low-confidence non-driver
Low-confidence driver
High-confidence driver
Num of mutations
BoostDM score
14.7%
5.7%
8.3%
71.3%
VAF
Fold increase of cases with CH cases
Age group
38-45
46-50
51-55
56-60
61-65
66-72
CH-Age associations
Odds Ratio (95% CI)

**Figure S7. Blood somatic mutations across UKB donors.**
a) VAF distribution of all mutations (prior to the application of boostDM-CH models) identified across the 12 CH driver genes. The broken line at 10% of VAF separates large clones and small clones used across the study.
b) From top to bottom, Mutational profile (tri-nucleotide context probabilities) of all potential CH mutations, boostDM-CH driver mutations and non-driver mutations in the UKB cohort and all observed blood somatic mutations from MSK-IMPACT cohort (representing neutral mutagenesis in HSCs). Notice the difference, particularly in the C>A channels.
c) Distribution of boostDM score for all UKB mutations, with vertical broken lines presenting an alternative (arbitrary) way of separating them into 4 tiers, instead of the driver/non-driver dichotomization. High-confidence driver (>=0.90), low-confidence driver (<0.9 and >=0.5), low-confidence non-driver (>0.1 and <0.5) and high-confidence non-driver (<=0.1).
d) VAF distribution of mutations in the 4 tiers, showing that the two categories of driver mutations concentrate at low VAF values.
e) Proportion of mutations identified in each gene across UKB donors that fall into the four tiers. The pie chart at the top of the figure represents the overall distribution of all observed mutations into these four tiers.
f) Left, fold increase of the number of cases with CH mutations in each of the 4 tiers of boostDM-CH scores (defined above) across age groups. Right, significance of the association of the presence of mutations in each tier with the age of donors, computed via logistic regression. Asterisks denote significant associations (p-value<0.05); ns: not significant. Associations with large positive effect size are only detected for mutations in tiers 3 and 4, although all associations tested are deemed significant.

### Figure S8

a

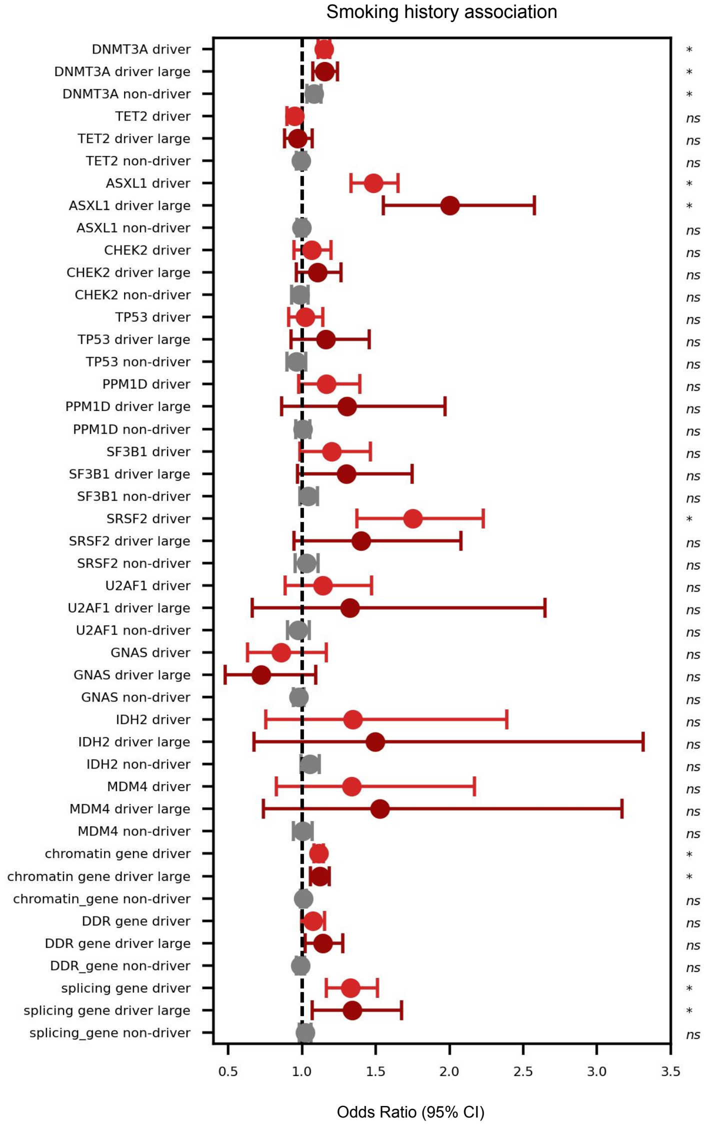
Smoking history association
DNMT3A driver
DNMT3A driver large
DNMT3A non-driver
TET2 driver
TET2 driver large
TET2 non-driver
ASXL1 driver
ASXL1 driver large
ASXL1 non-driver
CHEK2 driver
CHEK2 driver large
CHEK2 non-driver
TP53 driver
TP53 driver large
TP53 non-driver
PPM1D driver
PPM1D driver large
PPM1D non-driver
SF3B1 driver
SF3B1 driver large
SF3B1 non-driver
SRSF2 driver
SRSF2 driver large
SRSF2 non-driver
U2AF1 driver
U2AF1 driver large
U2AF1 non-driver
GNAS driver
GNAS driver large
GNAS non-driver
IDH2 driver
IDH2 driver large
IDH2 non-driver
MDM4 driver
MDM4 driver large
MDM4 non-driver
chromatin gene driver
chromatin gene driver large
chromatin_gene non-driver
DDR gene driver
DDR gene driver large
DDR_gene non-driver
splicing gene driver
splicing gene driver large
splicing_gene non-driver
Odds Ratio (95% CI)

b

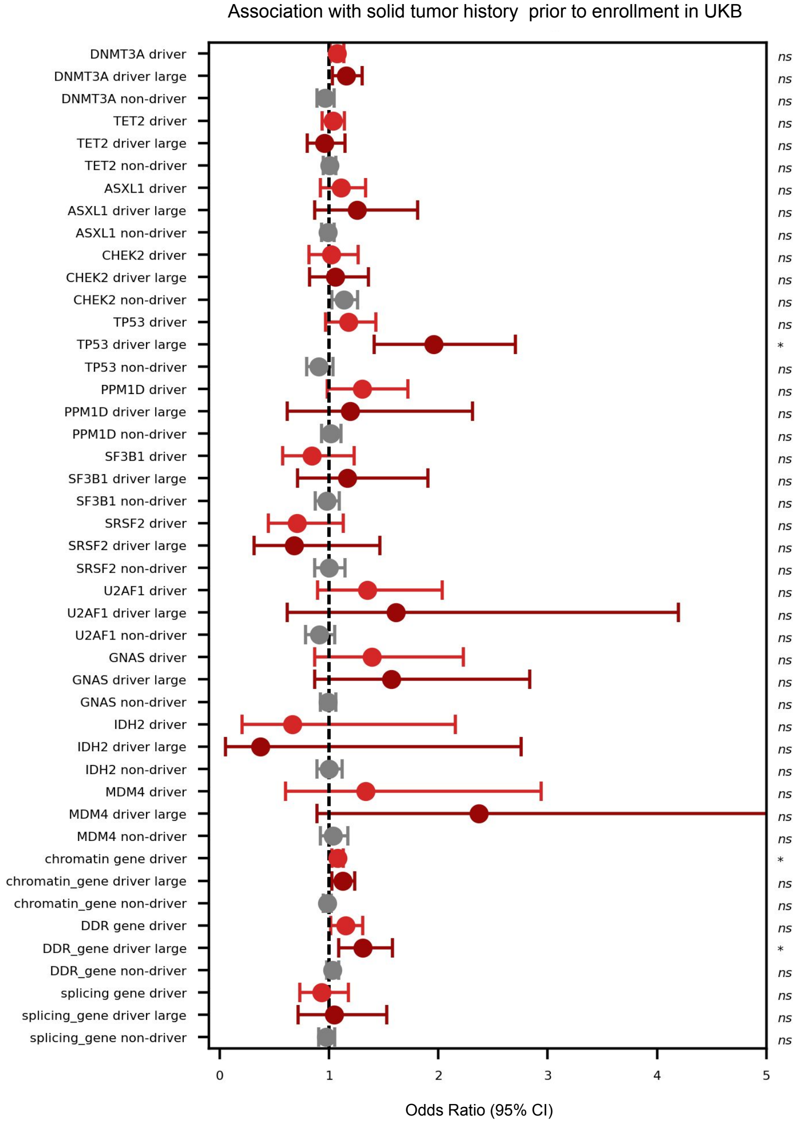
Association with solid tumor history prior to enrollment in UKB
DNMT3A driver
DNMT3A driver large
DNMT3A non-driver
TET2 driver
TET2 driver large
TET2 non-driver
ASXL1 driver
ASXL1 driver large
ASXL1 non-driver
CHEK2 driver
CHEK2 driver large
CHEK2 non-driver
TP53 driver
TP53 driver large
TP53 non-driver
PPM1D driver
PPM1D driver large
PPM1D non-driver
SF3B1 driver
SF3B1 driver large
SF3B1 non-driver
SRSF2 driver
SRSF2 driver large
SRSF2 non-driver
U2AF1 driver
U2AF1 driver large
U2AF1 non-driver
GNAS driver
GNAS driver large
GNAS non-driver
IDH2 driver
IDH2 driver large
IDH2 non-driver
MDM4 driver
MDM4 driver large
MDM4 non-driver
chromatin gene driver
chromatin_gene driver large
chromatin_gene non-driver
DDR gene driver
DDR_gene driver large
DDR_gene non-driver
splicing gene driver
splicing_gene driver large
splicing_gene non-driver
Odds Ratio (95% CI)

**Figure S8. CH associations with causal factors in the UKB.**

Association of the presence of CH driver and non-driver mutations (as predicted by the boostdm-CH models) with exogenous exposures known to be associated with this condition, namely smoking (a) and clinical history of a solid malignancy prior to enrollment in UKB, used as a proxy of cytotoxic treatment (b). Associations are computed via logistic regressions and are evaluated per gene and per functional gene group. Asterisks represent significant associations (FDR<0.05); ns: not significant.

### Figure S9

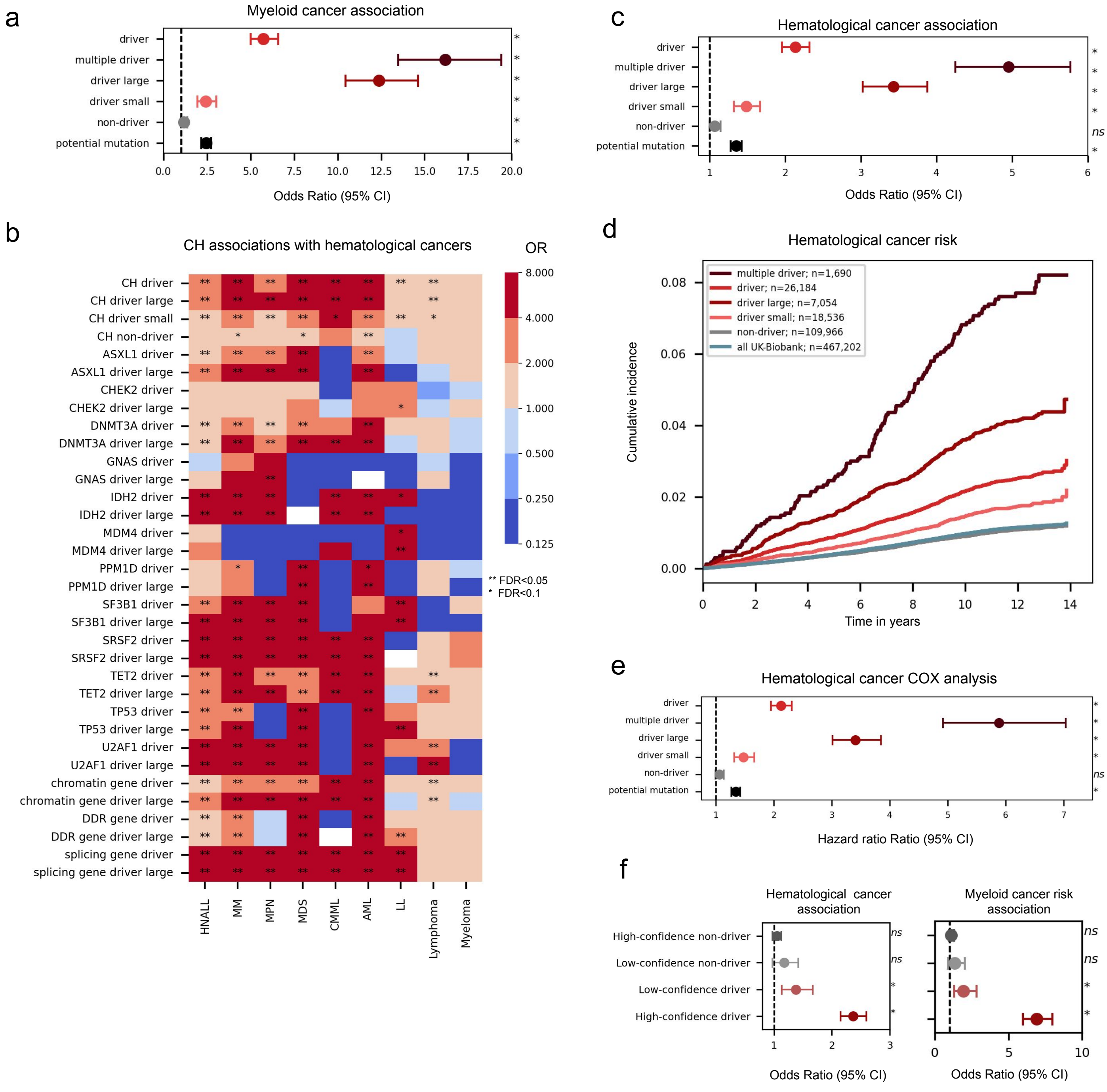
a
Myeloid cancer association
driver
multiple driver
driver large
driver small
non-driver
potential mutation
Odds Ratio (95% CI)
b
CH associations with hematological cancers
OR
CH driver
CH driver large
CH driver small
CH non-driver
ASXL1 driver
ASXL1 driver large
CHEK2 driver
CHEK2 driver large
DNMT3A driver
DNMT3A driver large
GNAS driver
GNAS driver large
IDH2 driver
IDH2 driver large
MDM4 driver
MDM4 driver large
PPM1D driver
PPM1D driver large
SF3B1 driver
SF3B1 driver large
SRSF2 driver
SRSF2 driver large
TET2 driver
TET2 driver large
TP53 driver
TP53 driver large
U2AF1 driver
U2AF1 driver large
chromatin gene driver
chromatin gene driver large
DDR gene driver
DDR gene driver large
splicing gene driver
splicing gene driver large
HNALL
MM
MPN
MDS
CMML
AML
LL
Lymphoma
Myeloma
** FDR<0.05
* FDR<0.1
c
Hematological cancer association
driver
multiple driver
driver large
driver small
non-driver
potential mutation
Odds Ratio (95% CI)
d
Hematological cancer risk
multiple driver; n=1,690
driver; n=26,184
driver large; n=7,054
driver small; n=18,536
non-driver; n=109,966
all UK-Biobank; n=467,202
Cumulative incidence
Time in years
e
Hematological cancer COX analysis
driver
multiple driver
driver large
driver small
non-driver
potential mutation
Hazard ratio Ratio (95% CI)
f
Hematological cancer association
Myeloid cancer risk association
High-confidence non-driver
Low-confidence non-driver
Low-confidence driver
High-confidence driver
Odds Ratio (95% CI)
Odds Ratio (95% CI)

**Figure S9. Associations between CH and the development of hematological cancers across UKB.**

a) Association between the presence of CH driver or non-driver mutations (as predicted by the boostdm-CH models) and the development of myeloid malignancies, computed via logistic regression in the UKB. The associations for large or small CH clones and clones with more than one CH driver mutation are also computed.

b) Association between the presence of CH drive mutations in individual genes or functional groups of genes and the development of different types of hematological cancers computed via logistic regression in UKB. The color of the cells represent the odds-ratio of the association, and the asterisks within each of them denote significant associations.

c,d,e) Association between the presence of CH driver (different groups) or non-driver mutations with the occurrence of hematological cancer measured via, logistic regression c), Kaplan-Meier analysis d), and Cox proportional hazards model e).

f) Association between either hematological or myeloid cancer and the presence of CH mutations grouped in four tiers according to their boostDM-CH scores (see supplementary figure S7), measured via logistic regression .

Asterisks represent significant associations (FDR<0.05); ns: not significant; HNALL: All hematological cancer types; MM: myeloid malignancies; MPN: Myeloproliferative neoplasms; MDS: Myelodysplastic syndromes; CMML: Chronic myelomonocytic leukemia; AML: Acute myeloid leukemias; LL: Lymphoid leukemias.

Figure S10

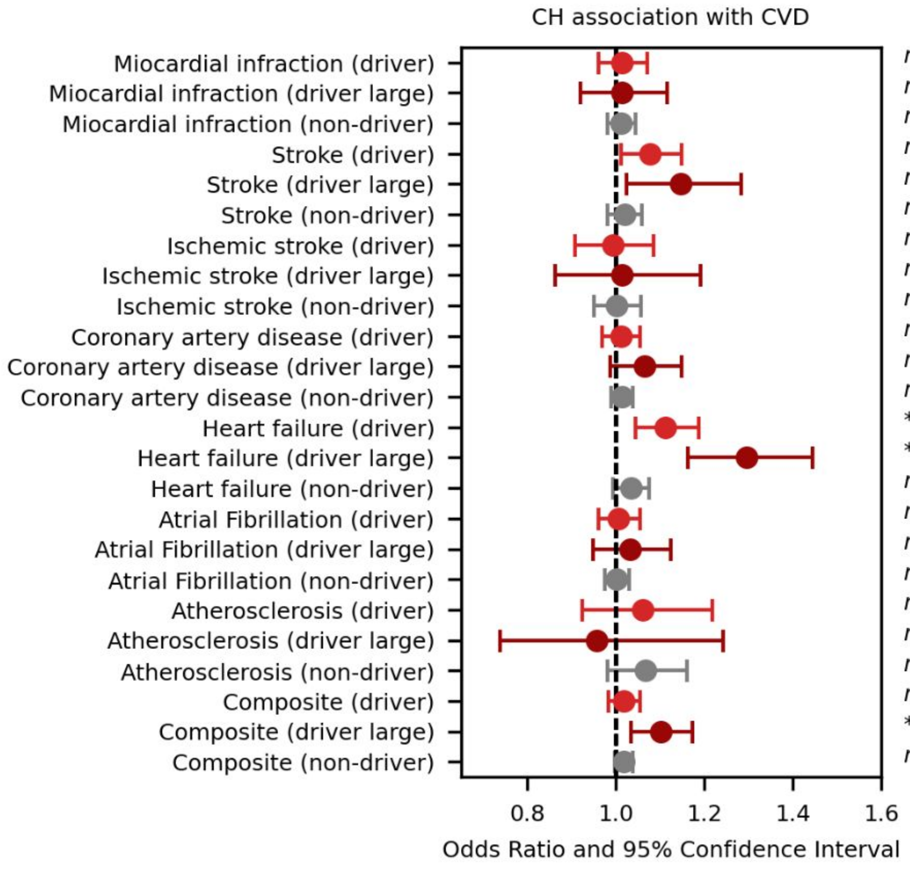
CH association with CVD
Miocardial infraction (driver)
Miocardial infraction (driver large)
Miocardial infraction (non-driver)
Stroke (driver)
Stroke (driver large)
Stroke (non-driver)
Ischemic stroke (driver)
Ischemic stroke (driver large)
Ischemic stroke (non-driver)
Coronary artery disease (driver)
Coronary artery disease (driver large)
Coronary artery disease (non-driver)
Heart failure (driver)
Heart failure (driver large)
Heart failure (non-driver)
Atrial Fibrillation (driver)
Atrial Fibrillation (driver large)
Atrial Fibrillation (non-driver)
Atherosclerosis (driver)
Atherosclerosis (driver large)
Atherosclerosis (non-driver)
Composite (driver)
Composite (driver large)
Composite (non-driver)
ns
ns
ns
ns
ns
ns
ns
ns
ns
ns
ns
ns
*
*
ns
ns
ns
ns
ns
ns
ns
ns
*
ns
0.8
1.0
1.2
1.4
1.6
Odds Ratio and 95% Confidence Interval

**Figure S10. Association of the presence of CH mutations with different cardiovascular events in UKB.**

The associations were measured using logistic regressions adjusted by several relevant covariables (see Methods). Asterisks represent significant associations (FDR<0.05); ns: not significant.

Figure S11

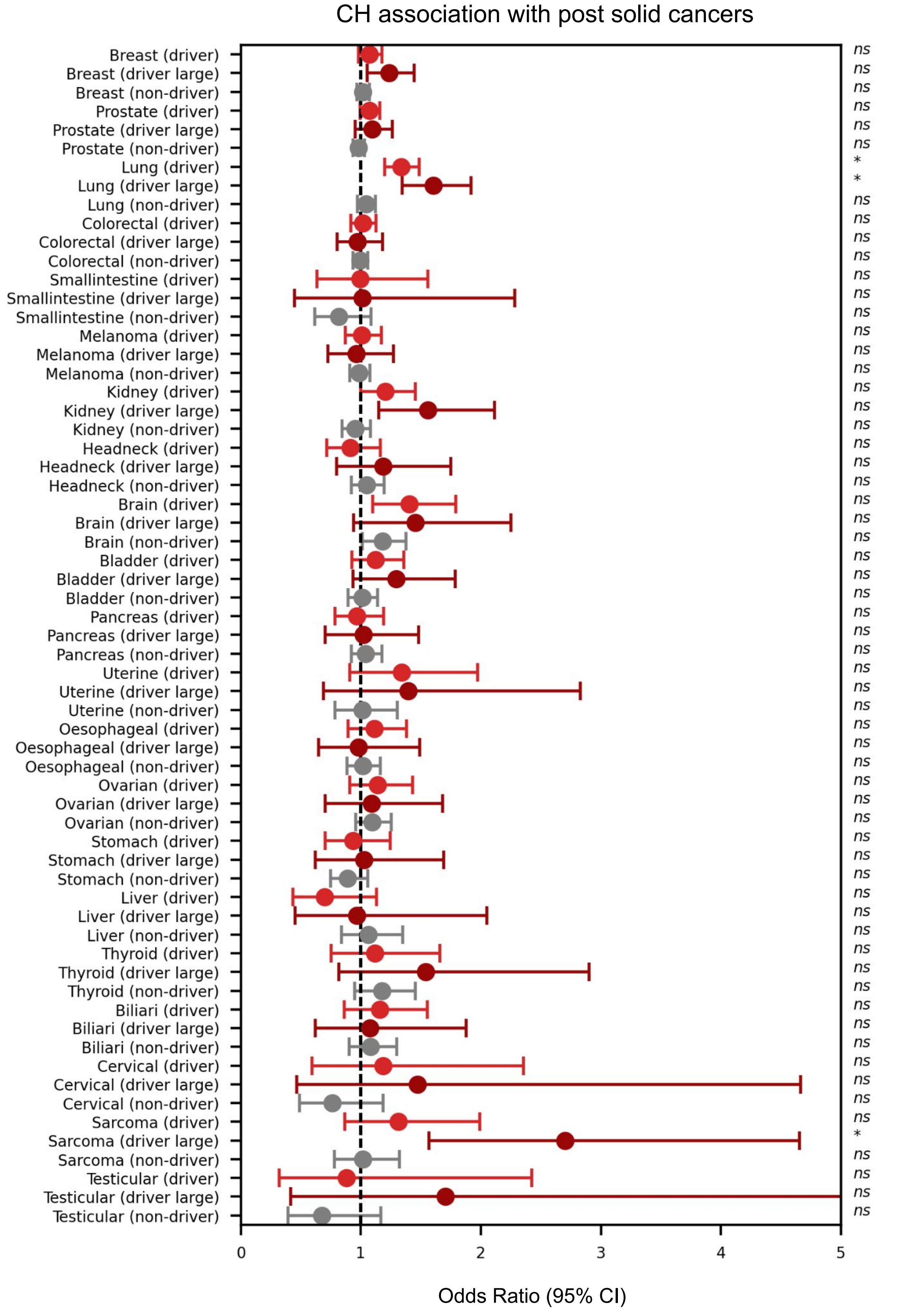
CH association with post solid cancers
Breast (driver) ns
Breast (driver large) ns
Breast (non-driver) ns
Prostate (driver) ns
Prostate (driver large) ns
Prostate (non-driver) ns
Lung (driver) *
Lung (driver large) *
Lung (non-driver) ns
Colorectal (driver) ns
Colorectal (driver large) ns
Colorectal (non-driver) ns
Smallintestine (driver) ns
Smallintestine (driver large) ns
Smallintestine (non-driver) ns
Melanoma (driver) ns
Melanoma (driver large) ns
Melanoma (non-driver) ns
Kidney (driver) ns
Kidney (driver large) ns
Kidney (non-driver) ns
Headneck (driver) ns
Headneck (driver large) ns
Headneck (non-driver) ns
Brain (driver) ns
Brain (driver large) ns
Brain (non-driver) ns
Bladder (driver) ns
Bladder (driver large) ns
Bladder (non-driver) ns
Pancreas (driver) ns
Pancreas (driver large) ns
Pancreas (non-driver) ns
Uterine (driver) ns
Uterine (driver large) ns
Uterine (non-driver) ns
Oesophageal (driver) ns
Oesophageal (driver large) ns
Oesophageal (non-driver) ns
Ovarian (driver) ns
Ovarian (driver large) ns
Ovarian (non-driver) ns
Stomach (driver) ns
Stomach (driver large) ns
Stomach (non-driver) ns
Liver (driver) ns
Liver (driver large) ns
Liver (non-driver) ns
Thyroid (driver) ns
Thyroid (driver large) ns
Thyroid (non-driver) ns
Biliari (driver) ns
Biliari (driver large) ns
Biliari (non-driver) ns
Cervical (driver) ns
Cervical (driver large) ns
Cervical (non-driver) ns
Sarcoma (driver) ns
Sarcoma (driver large) *
Sarcoma (non-driver) ns
Testicular (driver) ns
Testicular (driver large) ns
Testicular (non-driver) ns
0
1
2
3
4
5
Odds Ratio (95% CI)

**Figure S11. Association between the presence of CH mutations and the development of a posterior solid malignancy in the UKB.**

The associations for the presence of all driver mutations, driver mutations in large clones, or non-driver mutations, and the diagnoses of different solid malignancies after enrollment were measured using logistic regressions. Asterisks represent significant associations (FDR<0.05); ns: not significant.

Figure S12

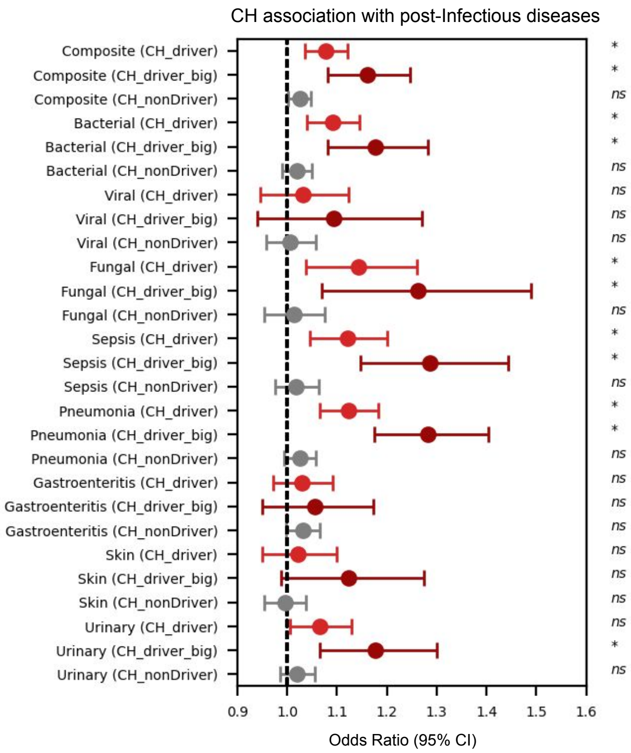
CH association with post-Infectious diseases
Composite (CH_driver)
Composite (CH_driver_big)
Composite (CH_nonDriver)
Bacterial (CH_driver)
Bacterial (CH_driver_big)
Bacterial (CH_nonDriver)
Viral (CH_driver)
Viral (CH_driver_big)
Viral (CH_nonDriver)
Fungal (CH_driver)
Fungal (CH_driver_big)
Fungal (CH_nonDriver)
Sepsis (CH_driver)
Sepsis (CH_driver_big)
Sepsis (CH_nonDriver)
Pneumonia (CH_driver)
Pneumonia (CH_driver_big)
Pneumonia (CH_nonDriver)
Gastroenteritis (CH_driver)
Gastroenteritis (CH_driver_big)
Gastroenteritis (CH_nonDriver)
Skin (CH_driver)
Skin (CH_driver_big)
Skin (CH_nonDriver)
Urinary (CH_driver)
Urinary (CH_driver_big)
Urinary (CH_nonDriver)
*
*
ns
*
*
ns
ns
ns
ns
*
*
ns
*
*
ns
*
*
ns
ns
ns
ns
ns
ns
ns
ns
*
ns
0.9
1.0
1.1
1.2
1.3
1.4
1.5
1.6
Odds Ratio (95% CI)

**Figure S12. CH associations with different infectious diseases in the UKB.**
Association of CH with infectious disease posteriorly to enrollment in the UKB, measured via logistic regression. Smoking history and occurrence of posterior hematological cancer were included as covariates. Asterisks represent significant associations (FDR<0.05); ns: not significant.

### Figure S13

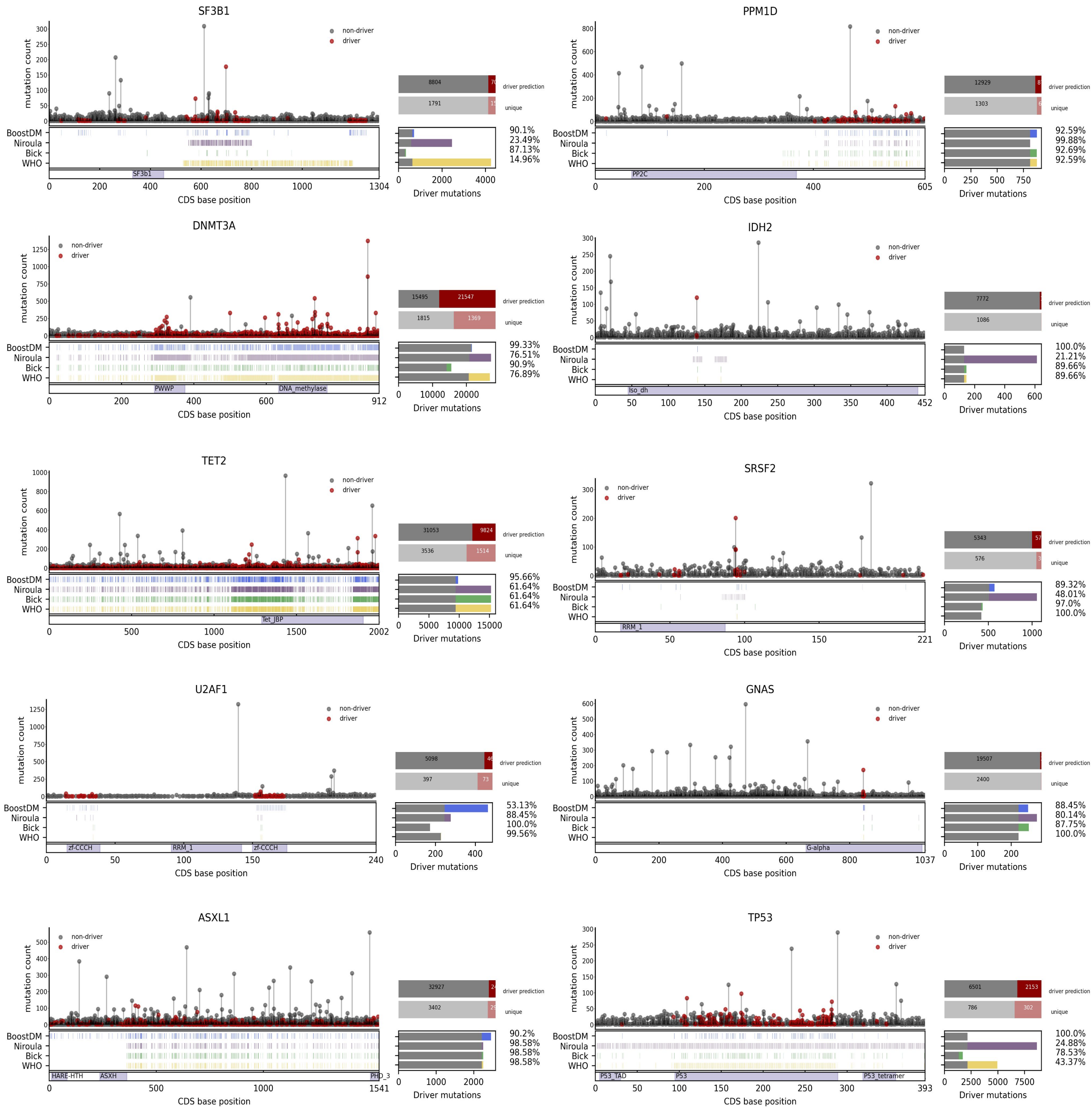

SF3B1
mutation count
non-driver
driver
8804
1791
driver prediction
unique
BoostDM
Niroula
Bick
WHO
SF3b1
CDS base position
Driver mutations
90.1%
23.49%
87.13%
14.96%
PPM1D
12929
1303
PP2C
92.59%
99.88%
92.69%
92.59%
DNMT3A
15495
21547
1815
1369
PWWP
DNA_methylase
99.33%
76.51%
90.9%
76.89%
IDH2
7772
1086
Iso_dh
100.0%
21.21%
89.66%
89.66%
TET2
31053
9824
3536
1514
Tet_JBP
95.66%
61.64%
61.64%
61.64%
SRSF2
5343
576
RRM_1
89.32%
48.01%
97.0%
100.0%
U2AF1
5098
397
73
zf-CCCH
RRM_1
53.13%
88.45%
100.0%
99.56%
GNAS
19507
2400
G-alpha
88.45%
80.14%
87.75%
100.0%
ASXL1
32927
3402
HARE-HTH
ASXH
PHD_3
90.2%
98.58%
98.58%
98.58%
TP53
6501
2153
786
302
P53_TAD
P53
P53_tetramer
100.0%
24.88%
78.53%
43.37%

**Figure S13. Comparison of boostDM-CH models and expert-curated rules in the identification of CH driver mutations in the UKB.**

Each panel represents a different CH gene. The needle plot at the top of each panel represents the distribution of mutations observed across UKB along the sequence of each protein, with driver mutations (according to boostDM-CH) colored red and non-driver mutations in gray. The height of each needle represents the frequency of mutations at a particular site of the protein. The bars at the right represent the proportion of driver (red) and non-driver (gray) mutations taking into account either all (top bar) or unique (bottom bar) mutations. The plot at the bottom of each panel presents the distribution of driver mutations according to the boostDM-CH model (top, blue), or any of the prior-knowledge rule sets introduced earlier (below, following the color code introduced in Fig. 1). The box at the right (analogously to Fig. S5), presents the proportion of mutations classified as drivers by boostDM-CH that are also classified as drivers by any of the expert-curated rule sets (top bar), or of mutations classified as drivers by one expert-curated rule set and boostDM-CH (other bars). In every bar, the gray segment represents the overlapping fraction, and the colored segments represent mutations uniquely classified by boostDM-CH or a particular rule set.

Figure S14

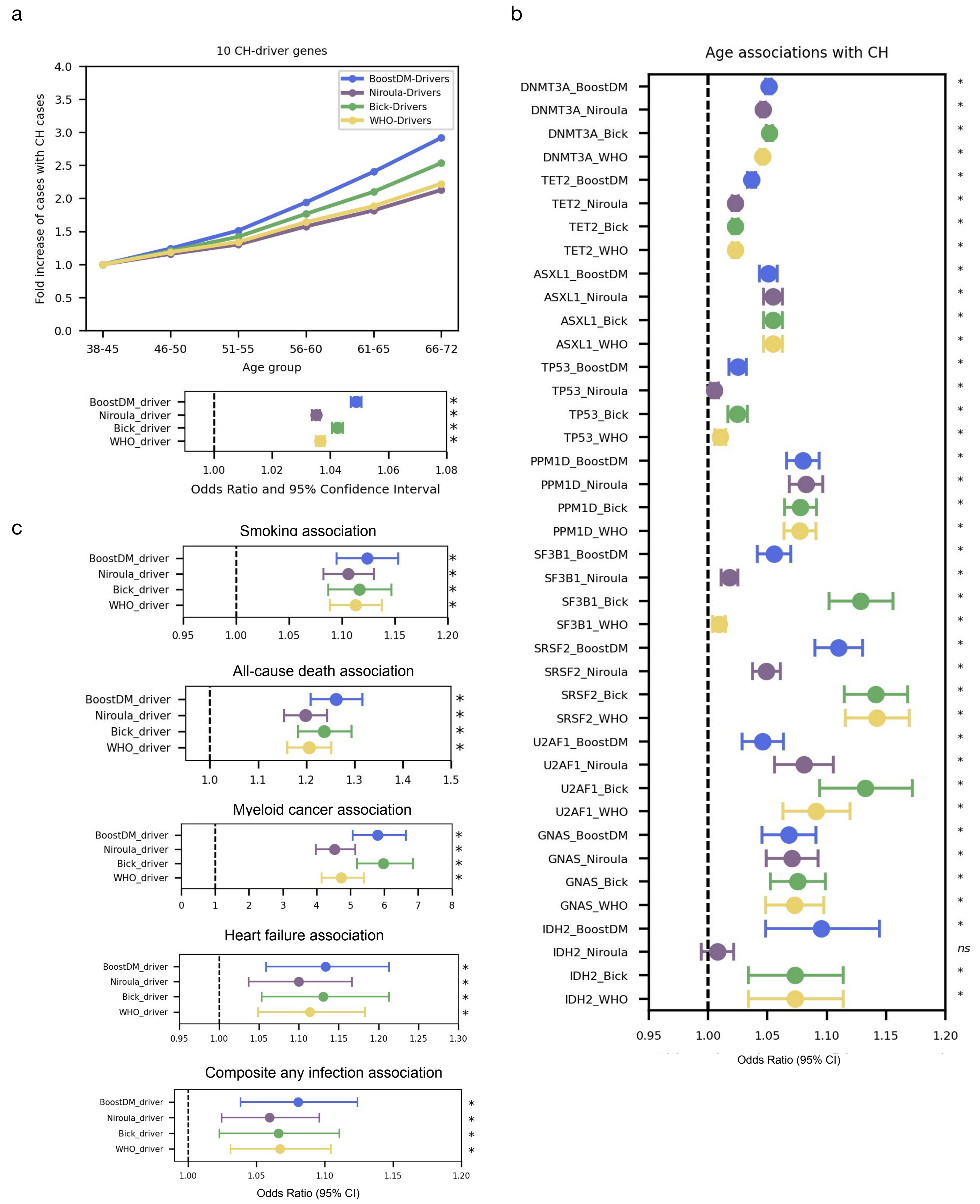

a
10 CH-driver genes
Fold increase of cases with CH cases
BoostDM-Drivers
Niroula-Drivers
Bick-Drivers
WHO-Drivers
38-45
46-50
51-55
56-60
61-65
66-72
Age group
BoostDM_driver
Niroula_driver
Bick_driver
WHO_driver
Odds Ratio and 95% Confidence Interval
b
Age associations with CH
DNMT3A_BoostDM
DNMT3A_Niroula
DNMT3A_Bick
DNMT3A_WHO
TET2_BoostDM
TET2_Niroula
TET2_Bick
TET2_WHO
ASXL1_BoostDM
ASXL1_Niroula
ASXL1_Bick
ASXL1_WHO
TP53_BoostDM
TP53_Niroula
TP53_Bick
TP53_WHO
PPM1D_BoostDM
PPM1D_Niroula
PPM1D_Bick
PPM1D_WHO
SF3B1_BoostDM
SF3B1_Niroula
SF3B1_Bick
SF3B1_WHO
SRSF2_BoostDM
SRSF2_Niroula
SRSF2_Bick
SRSF2_WHO
U2AF1_BoostDM
U2AF1_Niroula
U2AF1_Bick
U2AF1_WHO
GNAS_BoostDM
GNAS_Niroula
GNAS_Bick
GNAS_WHO
IDH2_BoostDM
IDH2_Niroula
IDH2_Bick
IDH2_WHO
ns
Odds Ratio (95% CI)
c
Smoking association
All-cause death association
Myeloid cancer association
Heart failure association
Composite any infection association
Odds Ratio (95% CI)

**Figure S14. Comparison of the association between CH driver mutations (following different approaches) and several phenotypes in the UKB.**

Results for the 10 genes included in the three sets of expert-curated rules introduced earlier are presented.

a) The top plot represents the fold increase in the number of cases with CH driver mutations across age groups. The driver mutations have been identified using either boostDM-CH models (blue) or the expert-curated rule sets introduced earlier (colors following the code introduced in Fig. 1). The bottom plot represents the strength of the association between age and the presence of CH driver mutations according to these different approaches, computed via logistic regression.

b) Association between the presence of CH driver mutations (according to the different approaches) in each of the 10 CH driver genes and age, computed through logistic regressions.

c) Association between the presence of CH driver mutations according to these different approaches and several clinical variables, measured via logistic regression. Asterisks represent significant associations (FDR<0.05); ns: not significant.

### Figure S15

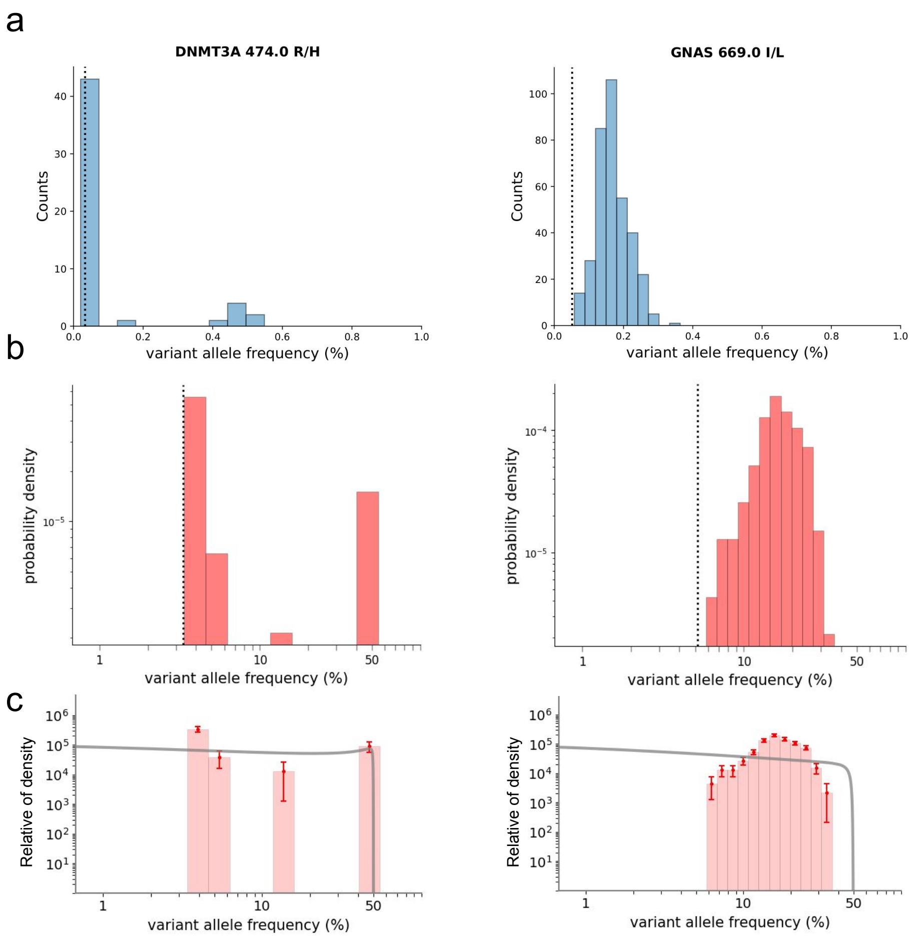

a
b
c
DNMT3A 474.0 R/H
GNAS 669.0 I/L
Counts
variant allele frequency (%)
probability density
Relative of density

**Figure S15. UKB VAF distribution of two variants excluded from the fitness estimation by visual inspection.**

The left half of the figure presents the VAF distribution across UKB samples of a SNV (*DNMT3A* R474H) with a bimodal distribution of VAF, which could point to the coexistence of a somatic and a germline variants across samples. The right half represents the VAF distribution of a SNV (*GNAS* I669L) that is potentially a germline variant.

a) VAF distribution of the two variants across UKB samples.

b) VAF distribution of the two variants across UKB samples in log scale.

c) Fitting of the relative density of the VAF distribution of both variants across UKB samples., which is used to estimate their fitness.

### Figure S16

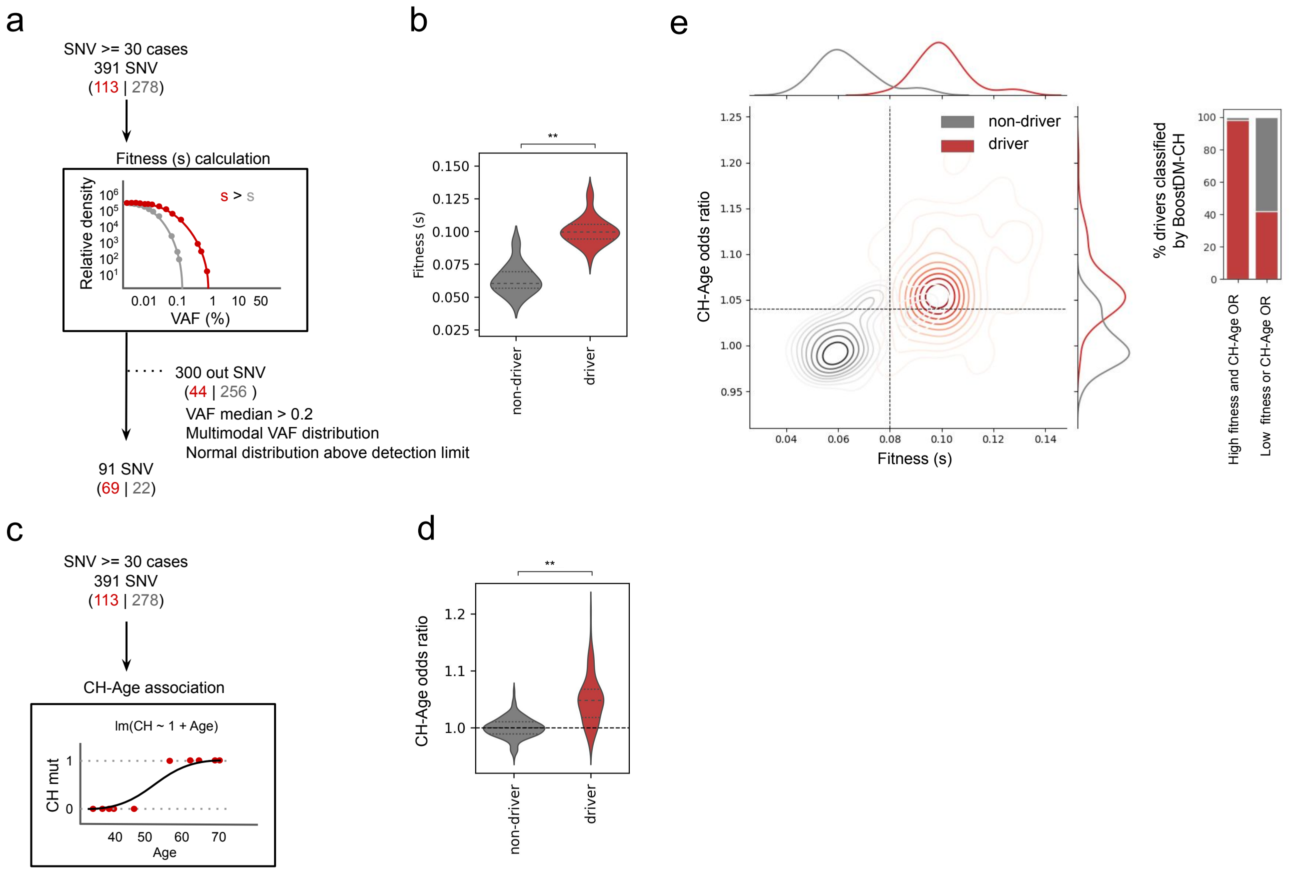

a
SNV >= 30 cases
391 SNV
(113 | 278)
Fitness (s) calculation
Relative density
s > s
VAF (%)
300 out SNV
(44 | 256 )
VAF median > 0.2
Multimodal VAF distribution
Normal distribution above detection limit
91 SNV
(69 | 22)
b
Fitness (s)
non-driver
driver
c
SNV >= 30 cases
391 SNV
(113 | 278)
CH-Age association
lm(CH ~ 1 + Age)
CH mut
Age
d
CH-Age odds ratio
non-driver
driver
e
non-driver
driver
CH-Age odds ratio
Fitness (s)
% drivers classified by BoostDM-CH
High fitness and CH-Age OR
Low fitness or CH-Age OR

**Figure S16. The fitness of recurrent blood somatic SNVs across 434,911 UKB donors.**
Same analysis as the one presented in Figure 5 of the main manuscript, excluding from the fitness estimation all UKB samples with more than one mutation in any of the 12 CH driver genes analyzed. In all, 34,969 samples with multiple mutations were excluded.
a) Schematic representation of the fitness score calculation for individual recurrent blood somatic SNVs, based on their observed VAF distribution across the UKB cohort.
b) Overall distribution of fitness scores of CH driver and non-driver mutations.
c) Left, schematic representation of the calculation of the associations (odds-ratio and p-value) between individual CH driver SNVs and age using logistic regression.
d) Distribution of odds-ratios computed for the association of driver and non-driver CH mutations with age (ageOR).
e) Left, kernel density estimate (KDE) representing the bi-dimensional density of SNVs in the ageOR- fitness score plane. The horizontal dashed line represents an ageOR of 1.04, the minimum rendering a significant p-value, whereas the vertical line is arbitrarily set at a fitness score of 0.08. These lines effectively separate the bi-dimensional distributions of driver and non-driver recurrent CH SNVs into four quadrants, with the density of both distributions clearly separated in the top-right (drivers) and bottom-left (non-drivers) quadrants. The distribution of fitness and ageOR for driver and non-driver mutations are presented along the x-axis and the y-axis of the plot, respectively. The proportions of driver and non-driver mutations in these two quadrants of the plot are represented in the stacked bars at the right of the plot.
In the figure, red denotes CH driver SNVs, whereas gray represents CH non-driver SNVs. Asterisks represent significant associations (FDR<0.05). Double asterisk represents p-value <0.001.

### Figure S17

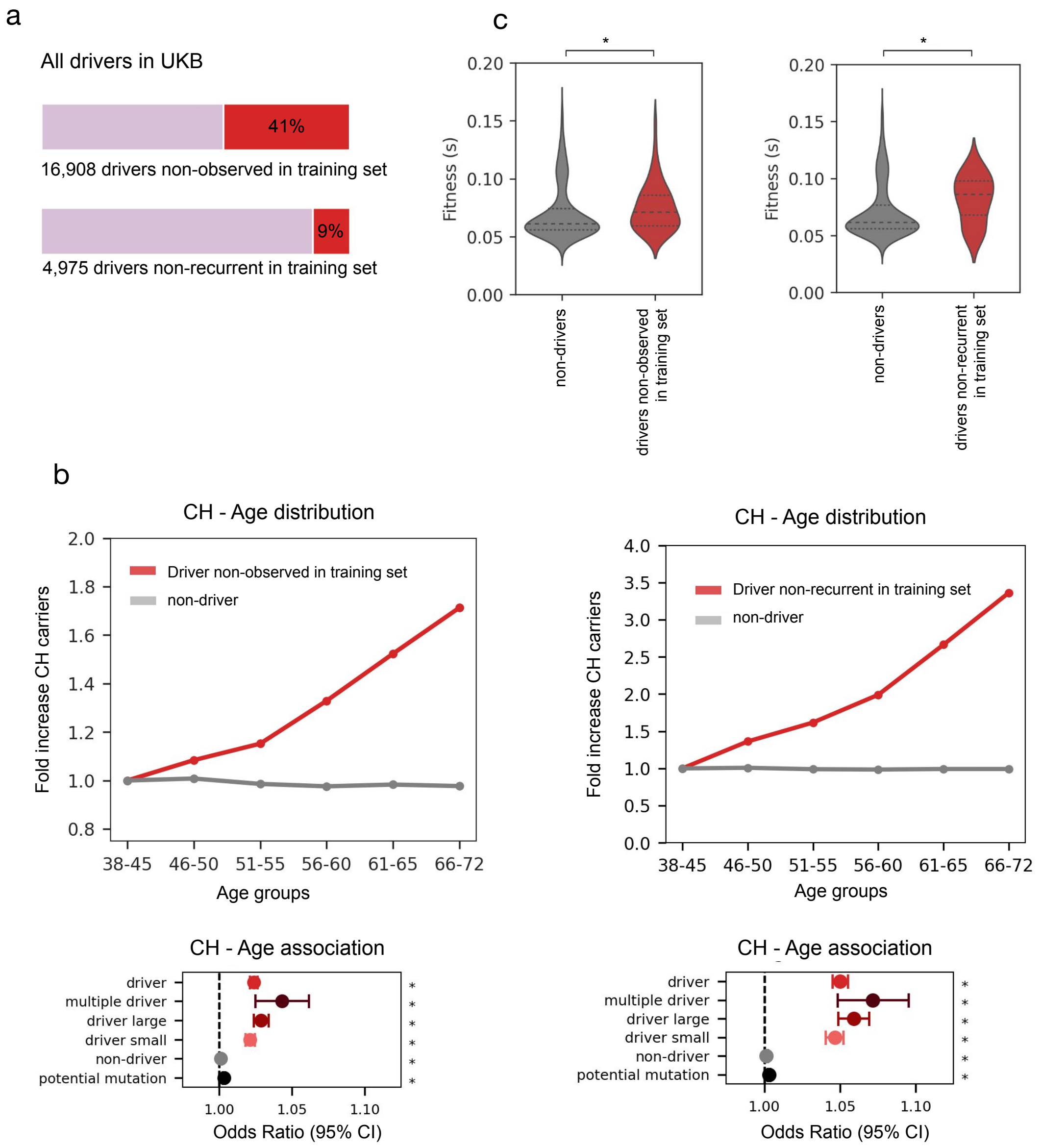
a
All drivers in UKB
41%
16,908 drivers non-observed in training set
9%
4,975 drivers non-recurrent in training set
c
Fitness (s)
non-drivers
drivers non-observed in training set
drivers non-recurrent in training set
b
CH - Age distribution
Driver non-observed in training set
non-driver
Driver non-recurrent in training set
Fold increase CH carriers
38-45
46-50
51-55
56-60
61-65
66-72
Age groups
CH - Age association
driver
multiple driver
driver large
driver small
non-driver
potential mutation
Odds Ratio (95% CI)

**Figure S17. UKB driver mutations that are not observed or not recurrent in the discovery cohorts.**

a) Proportion of driver mutations in UKB that are observed 0 (top) or 1 (bottom) times in the discovery cohorts.

b) Fold increase of the number of cases in UKB with CH driver mutations observed 0 (left) or 1 (right) times in the discovery cohorts. The bottom plots present the association of the presence of CH driver mutations in these two categories with age (logistic regression).

c) Overall distribution of fitness scores of 57 CH driver mutations observed 0 (left) or 1 (right) times in the discovery cohorts, in comparison to that of all non-driver recurrent mutations observed in UKB.

### Supplementary Tables

**Table S1. CH driver mutations expert-curated rules.**

| Gene name | Transcript | Bick | Niroula | WHO |
|---|---|---|---|---|
| ASXL1 | NM_015338 | Frameshift/nonsense/splice-site in exon 11-12 | Frameshift/stop gain/splice site in exon 11-12 | Frameshift/nonsense/splice-site in exon 11-12 |
| CHEK2 | | | Frameshift, nonsense, splice site | |

| | | | | |
|---|---|---|---|---|
| DNMT3A | NM_022552 | Frameshift/nonsense/splice-site, F290I, F290C, V296M, P307S, P307R, R326H, R326L, R326C, R326S, G332R, G332E, V339A, V339M, V339G, L344Q, L344P, R366P, R366H, R366G, A368T, A368V, R379H, R379C, I407T, I407N, I407S, F414L, F414S, F414C, A462V, K468R, C497G, C497Y, Q527H, Q527P, Y533C, S535F, C537G, C537R, G543A, G543S, G543C, L547H, L547P, L547F, M548I, M548K, G550R, W581R, W581G, W581C, R604Q, R604W, R635W, R635Q, S638F, G646V, G646E, L653W, L653F, I655N, V657A, V657M, R659H, Y660C, V665G, V665L, M674V, R676W, R676Q, G685R, G685E, G685A, D686Y, D686G, R688H, G699R, G699S, G699D, P700L, P700S, P700R, P700Q, P700T, P700A, D702N, D702Y, V704M, V704G, I705F, I705T, I705S, I705N, G707D, G707V, C710S, C710Y, S714C, V716D, V716F, V716I, N717S, N717I, P718L, R720H, R720G, K721R, K721T, Y724C, R729Q, R729W, R729G, F731C, F731L, F731Y, F731I, F732del, F732C, F732S, F732L, E733G, E733A, F734L, F734C, Y735C, Y735N, Y735S, R736H, R736C, R736P, L737H, L737V, L737F, L737R, A741V, P742P, P743R, P743L, R749C, R749L, R749H, R749G, F751L, F751C, F752del, F752C, F752L, F752I, F752V, W753G, W753C, W753R, L754P, L754R, L754H, F755S, F755I, F755L, M761I, M761V, G762C, V763I, S770L, S770W, S770P, R771Q, F772I, F772V, L773R, L773V, E774K, E774D, E774G, I780T, D781G, R792H, W795C, W795L, G796D, G796V, N797Y, N797H, N797S, P799S, P799R, P799H, R803S, R803W, P804L, P804S, K826R, S828N, K829R, T835M, N838D, K841Q, Q842E, P849L, D857N, W860R, E863D, F868S, G869S, G869V, M880V, S881R, S881I, R882H, R882P, R882C, R882G, A884P, A884V, Q886R, L889P, L889R, G890D, G890R, G890S, V895M, P896L, V897G, V897D, R899L, R899H, R899C, L901R, L901H, P904L, F909C, P904Q, A910P, C911R, C911Y | Frameshift/stop gain/splice site, Nonsynonymous AA290-390, AA547-911, AA414, AA494, AA497, AA508, AA527, AA529, AA531, AA532, AA537, AA543 | Frameshift/nonsense/splice-site; Missense in aa range: p.292-350, p.482-614 and p.634-912 |

| | | | | |
|---|---|---|---|---|
| GNAS | NM_000516 | R201S, R201C, R201H, R201L, Q227K, Q227R, Q227L, Q227H, R374C | Nonsynonymous AA201, AA227, AA844 | R201 |
| IDH2 | NM_002168 | R140W, R140Q, R140L, R140G, R172W, R172G, R172K, R172T, R172M, R172N, R172S | Nonsynonymous AA134-146, 164-180 | R140, R172 |
| PPM1D | NM_003620 | Frameshift/nonsense, exon 5 or 6 | Frameshift/stop gain/splice site AA421-605 | Frameshift/nonsense/splice-site in exon 5/6 |
| SF3B1 | NM_012433 | G347V, R387W, R387Q, E592K, E622D, Y623C, R625L, R625C, R625G, H662Q, H662D, T663I, K666N, K666T, K666E, K666R, K700E, V701F, A708T, G740R, G740E, A744P, D781G, E783K, R831Q, L833F, E862K, R957Q | Nonsynonymous AA550-800 | Missense in terminal HEAT domains (p.529-1201) |
| SRSF2 | NM_003016 | Y44H, P95H, P95L, P95T, P95R, P95A, P107H, P95fs | Nonsynonymous AA57, AA95, AA85-100 | Missense/in-frame deletion involving P95 |
| TET2 | NM_001127208 | Frameshift/nonsense/splice-site, missense mutations in catalytic domains (p.1104-1481 and 1843-2002) | Frameshift/stop gain/splice site, Nonsynonymous AA1104-1481, AA1843-2002 | Frameshift/nonsense/splice-site; Missense in aa range: p.1104-1481 and p.1843-2002 |

| | | | | |
|---|---|---|---|---|
| TP53 | NM_001126112 | Frameshift/nonsense/splice-site, S46F, G105C, G105R, G105D, G108S, G108C, R110L, R110C, T118A, T118R, T118I, S127F, S127Y, L130V, L130F, K132Q, K132E, K132W, K132R, K132M, K132N, F134V. F134L, F134S, C135W, C135S, C135F, C135G, C135Y, Q136K, Q136E, Q136P, Q136R, Q136L, Q136H, A138P, A138V, A138A, A138T, T140I, C141R, C141G, C141A, C141Y, C141S, C141F, C141W, V143M, V143A, V143E, L145Q, W146C, W146L, L145R, V147G, P151T, P151A, P151S, P151H, P151R, P152S, P152R, P152L, T155P, T155A, V157F, R158H, R158L, A159V, A159P, A159S, A159D, A161T, A161D, Y163N, Y163H, Y163D, Y163S, Y163C, K164E, K164M, K164N, K164P, H168Y, H168P, H168R, H168L, H168Q, M169I, M169T, M169V, E171K, E171Q, E171G, E171A, E171V, E171D, V172D, V173M, V173L, V173G, R174W, R175G, R175C, R175H, C176R, C176G, C176Y, C176F, C176S, P177R, P177R, P177L, H178D, H178P, H178Q, H179Y, H179R, H179Q, R181C, R181Y, D186G, G187S, P190L, P190T, H193N, H193P, H193L, H193R, L194F, L194R, I195F, I195N, I195T, R196P, V197L, G199V, Y205N, Y205C, Y205H, D208V, R213Q, R213P, R213L, R213Q, H214D, H214R, S215G, S215I, S215R, V216M, V217G, Y220N, Y220H, Y220S, Y220C, E224D, I232F, I232N, I232T, I232S, Y234N, Y234H, Y234S, Y234C, Y236N, Y236H, Y236C, M237V, M237K, M237I, C238R, C238G, C238Y, C238W, N239T, N239S, S241Y, S241C, S241F, C242G, C242Y, C242S, C242F, G244S, G244C, G244D, G245S, G245R, G245C, G245D, G245A, G245V, G245S, M246V, M246K, M246R, M246I, N247I, R248W, R248G, R248Q, R249G, R249W, R249T, R249M, P250L, I251N, L252P, I254S, I255F, I255N, I255S, L257Q, L257P, E258K, E258Q, D259Y, S261T, G262D, G262V, L265P, G266R, G266E, G266V, R267W, R267Q, R267P, E271K, V272M, V272L, R273S, R273G, R273C, R273H, R273P, R273L, V274F, V274D, V274A, V274G, V274L, C275Y, C275S, C275F, A276P, C277F, C277Y, P278T, P278A, P278S, P278H, P278R, P278L, G279E, R280G, R280K, R280T, R280I, R280S, D281N, D281H, D281Y, D281G, D281E, R282G, R282W, R282Q, R282P, E285K, E285V, E286G, E286V, E286K, K320N, L330R, G334V, R337C, R337L, A347T, L348F, T377P | Frameshift/stop gain/splice site, Nonsynonymous | Frameshift/nonsense/splice-site; Missense in aa range: p.72, p.95-288 and p.337 |

| | | | | |
|---|---|---|---|---|
| U2AF1 | NM_006758 | D14G, S34F, S34Y, R35L, R156H, R156Q, Q157R, Q157P | Nonsynonymous AA22, AA28, AA32, AA34, AA154, AA156, AA157 | Missense at p.S34 / p.R156 / p.Q157 |

**Table S2. Cancer and disease definitions by UKB.** Definitions defined by using ICD-10/ICD-9, self-reported cancer, self-reported non-cancer illness, operation and OPSC4.

| Disease category | Disease outcome | Abbreviation | Sex specific | ICD-10 codes | ICD-9 codes | Cancer codes, self-reported | Non-cancer illness codes, self-reported | Operation code | OPCS4 codes |
|---|---|---|---|---|---|---|---|---|---|
| HNALL | Myeloid malignancies | MM | | C92.0, C92.2-9, C93.0-2, C93.7, C93.9, C94.0, C94.2-6, C96.2, D45, D46, D47.0-1, D47.3-5 | 205.0, 205.2, 205.3, 205.8, 205.9, 206.0, 206.2, 207.0, 207.2, 238.4, 238.5, 238.7 | 1051, 1074 | 1449, 1438 | | |
| HNALL | Myeloproliferative neoplasms | MPN | | D45, D47.0, D47.1, D47.3, D47.4 | 238.4, 238.5 | | 1449, 1438 | | |
| HNALL | Myelodysplastic syndromes | MDS | | C946, D46 | 238.7 | 1051 | | | |
| HNALL | Chronic myelomonocytic leukemia | CMML | | C93.1 | 206.1 | | | | |
| HNALL | Acute myeloid leukemias | AML | | C92.0, C92.2-9, C93.0, C93.2, C94.0, C94.2 | 205.0, 205.2, 205.3, 205.8, 205.9, 206.0, 206.2, 207.0, 207.2 | 1074 | | | |
| HNALL | Lymphoid leukemias | LL | | C91 | 204 | 1055 | | | |
| HNALL | Lymphoma | | | C81-88 | 200-202 | 1047, 1052, 1053 | | | |
| HNALL | Myeloma | | | C90.0, C90.1 | 203.0, 203.1 | 1050 | | | |
| Cancer | Breast | | F | C50, Z853 | 174, 175, V103 | 1022 | | | |
| Cancer | Prostate | | M | C61 | 185 | 1044 | | | |

| | | | | | | |
|---|---|---|---|---|---|---|
| Cancer | Lung | | | C33, C34, C399, Z851 | 162, V101 | 1001, 1027, 1028, 1080 |
| Cancer | Colorectal | | | C18-C21 | 153, 154 | 1020-1023 |
| Cancer | Small intestine | | | C17 | 152 | 1019 |
| Cancer | Melanoma | | | C43 | 172 | 1059 |
| Cancer | Kidney | | | C64, Z855 | 189.0, V105 | 1034 |
| Cancer | Head and neck | | | C00-C14, C30-C32 | 140-149, 160, 161 | 1006, 1007, 1009, 1004, 1010, 1011, 1012, 1077, 1078, 1079, 1005, 1015, 1016 |
| Cancer | Brain | | | C70, C71, C72.0, C72.3 | 191, 192.0, 192.1, 192.2, 192.3 | 1031, 1032, 1033 |
| Cancer | Bladder | | | C65, C66, C67, C723 | 188, 189.1, 189.2 | 1035 |
| Cancer | Pancreas | | | C25 | 157 | 1026 |
| Cancer | Uterine | | F | C54, C55 | 179, 182 | 1040 |
| Cancer | Oesophageal | | | C15 | 150 | 1017 |
| Cancer | Ovarian | | F | C56, C57.0, C57.4 | 183.0, 183.2, 183.8, 183.9 | 1039, 1087 |
| Cancer | Stomach | | | C16, Z85.02 | 151 | 1018 |
| Cancer | Liver | | | C22.0 | 155.0 | 1024 |
| Cancer | Thyroid | | | C73 | 193 | 1065 |

| | | | | | | | | | |
|---|---|---|---|---|---|---|---|---|---|
| Cancer | Biliari | | | C23, C24, C22.1 | 155.1, 156.0 | 1025 | | | |
| Cancer | Cervical | | F | C53 | 180 | 1041 | | | |
| Cancer | Sarcoma | | | C49 | 171 | 1068 | | | |
| Cancer | Testicular | | M | C62 | 186 | 1045 | | | |
| CVD | Myocardial infarction | MI | | I21, I22, I23, I24, 125.2 | 410, 411.0, 411.9 | | 1075 | | |
| CVD | Stroke | | | I60, I61, I63, I64 | 430, 431, 434, 436 | | 1081, 1086, 1491, 1583 | | |
| CVD | Ischemic stroke | | | I63 | 434, 436 | | 1583 | | |
| CVD | Coronary artery disease | CAD | | I21, I22, I23, I24, I25.1, I25.2, I25.5, I25.6, I25.8, I25.9 | 410, 411, 412, 414.0, 414.8, 414.9 | | 1075 | 1070, 1095, 1523 | K40-K46, K49, K50.1, K50.2, K50.4, K75 |
| CVD | Heart failure | HF | | I11.0, I13.0, I13.2, I50 | 428 | | 1076 | | |
| CVD | Atrial fibrillation | AF | | I48 | 4273 | | 1471, 1483 | | |
| CVD | Atherosclerosis | ATH | | I70 | 440 | | | | |
| Other | Type-2 diabetes mellitus | T2DM | | E11, E13, E14 | 250 | | 1223, 1220 | | |
| Other | Hypertension | HYP | | I10-I13, I15 | 401-405 | | 1065, 1072 | | |
